# Plant based enteral nutrition outperforms artificial nutrition in mitigating consequences of antibiotic-induced dysbiosis in mice and humans

**DOI:** 10.1101/2025.03.19.25323813

**Authors:** Mona Chatrizeh, Jianmin Tian, Matthew Rogers, Firuz Feturi, Guojun Wu, Brian Firek, Roman Nikonov, Lauren Cass, Alexandra Sheppeck, Rafael G. Ramos-Jiménez, Lavnish Ohja, Ali Caroll, Mathew Henkel, Justin Azar, Rajesh K. Aneja, Brian Campfield, Dennis Simon, Michael J. Morowitz

## Abstract

Malnutrition, gut inflammation, and antibiotic induced dysbiosis (AID) are omnipresent risk factors for poor clinical outcomes among critically ill patients. We previously showed that commercially available plant-based enteral nutrition (PBEN) preserves a commensal microbiome when compared to commonly used forms of commercially available artificial enteral nutrition (AEN). This study reveals that PBEN is superior to artificial enteral nutrition (AEN) in recovering from antibiotic-induced dysbiosis (AID) in mice and humans. PBEN effectively mitigates anemia, leukopenia, restores naïve lymphocyte populations, and reduces bone marrow myeloid cell expansion. Animals randomized to PBEN also fared better in response to infectious challenges after antibiotics. A pilot clinical study validated these findings, showing increased gut commensals, reduced pathogens, and improved leukocyte balance in critically ill patients receiving PBEN compared to AEN. These results suggest PBEN offers a practical dietary approach to mitigate antibiotic-associated complications and improve clinical outcomes among hospitalized patients requiring supplemental nutrition.

## Introduction

Critically ill patients exhibit deranged microbiomes due to various factors, including the prevalent use of antibiotics (ABX)^1^. While antibiotic use in the ICU is understandable given the severity of illness, ABX treatment is associated with poor outcomes beyond the well-known risk of antimicrobial resistance^2^. In fact, ABX are more likely than any other class of drugs to be associated with adverse drug events (ADEs)^3^ including GI disturbances such as *C. difficile* infection, hematologic complications, and increased length of hospital stay, cost of care, and risk of mortality^3,4^. Even in the absence of detectable ADEs, ABX treatment is associated with poor outcomes in specific clinical contexts such as readmission for sepsis within 90d of hospital discharge^5^, and complications after elective surgery^6^. In addition, researchers have reported increased mortality and morbidity following ABX depletion of the microbiome in a range of animal models including bacterial pneumonia^7^, fungal sepsis^8^, and experimental pancreatitis^9^. The underlying mechanisms for these complications are not fully understood, but it is clear that eradicating the gut microbiome with ABX can be harmful.

Despite improved stewardship programs, antimicrobial usage during critical illness is likely to remain prevalent. The utilization of antibiotics in critical care underscores the need for strategies to rescue or optimize the gut microbiome. Current approaches include probiotic administration^10^ and fecal microbiota transplantation^11^, but adoption of these approaches is limited due to context-specific safety concerns. Dietary interventions are a less explored strategy for mitigating ICU dysbiosis. Emerging evidence suggests that dietary composition can modify both antibiotic efficacy and the unintended consequences of antibiotic use^12^. In particular, increasing nutritional fiber intake shows great promise^13^ particularly in immunotherapy ^14^ but few studies have investigated the impact of dietary interventions on antibiotic treatment response in humans^13,15^.

The most common form of supplemental nutrition for patients unable to eat by mouth consists of commercially available liquid formulas containing prespecified amounts of macro and micronutrients, which we refer to as conventional or artificial enteral nutrition (AEN). These chemically defined formulas are notable for low fiber and high sugar content, and they contain preservatives and emulsifiers with pro-inflammatory properties^16,17^. More recently, several forms of all-natural plant-based enteral nutrition (PBEN) have been released commercially. These formulas are high in dietary fiber and low in added sugar, but their efficacy compared to traditional AEN has not been systematically evaluated. We and others have shown that PBEN is well tolerated in humans and prevents gut inflammation in murine models by promoting the growth of commensal anaerobes ^18,19^.

This study aims to compare the effects of AEN and PBEN on antibiotic-induced dysbiosis (AID). Murine models were used to evaluate the impact of these diets on the gut microbiota, immune cell homeostasis, and susceptibility to infection after exposure to broad spectrum antibiotics. Rather than studying experimental rodent formulas to study individual dietary components such as fiber or glucose, we chose to directly examine the performance of intact commercially available human formulas in our murine model as others have done^20–23^. Additionally, we conducted a single-center trial involving critically ill children randomized to either AEN or PBEN to assess the clinical implications of enteral formula choice on the gut microbiome. Unexpectedly, we observed an association between enteral diet and lymphopenia, a clinical predictor of survival.

## Results

### PBEN mitigates antibiotic-induced dysbiosis in mice

The composition and function of the microbiome is intrinsically shaped by use of antibiotics and choice of diet. We utilized 16S rRNA gene sequencing to monitor temporal variations in response to antibiotic and dietary interventions. Mice received a cocktail of broad-spectrum oral antibiotics (AVNM) for seven days, followed by a recovery period during which they were randomized to receive either plant-based enteral nutrition (PBEN) or artificial enteral nutrition (AEN) for an additional seven days. Sequencing data revealed a significant decline in alpha diversity (Chao1) in all antibiotic-exposed animals, indicating a reduction in microbial richness and diversity (Figure 1B). However, PBEN randomized animals displayed a rebound following ABX cessation whereas AEN randomized animals sustained a decrease in diversity. Surprisingly, ABX *naive* animals fed with AEN (but not PBEN) also experienced a collapse in alpha diversity similar to ABx treated animals.

**Figure 1.**
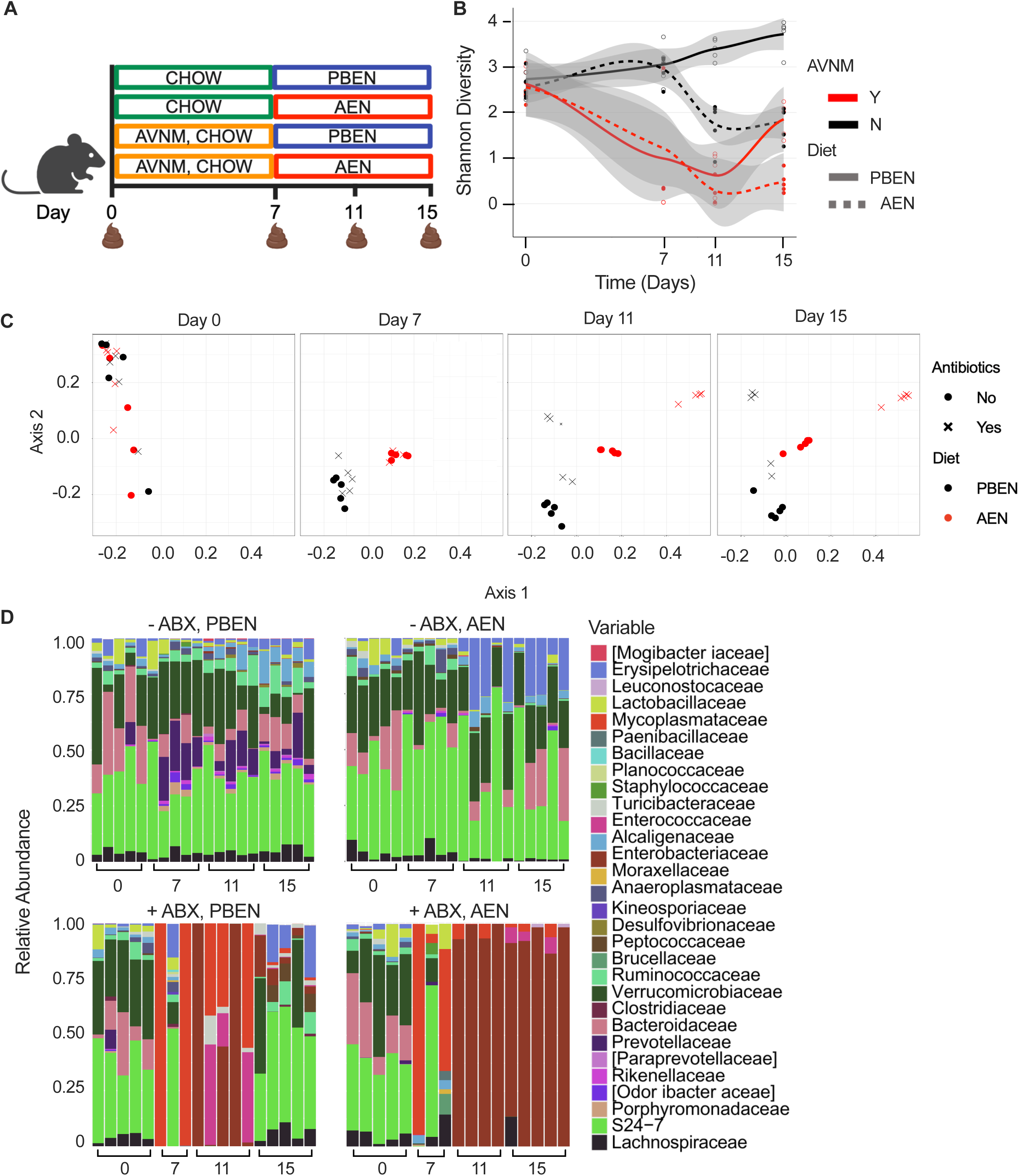
PBEN is superior to AEN in restoring microbial diversity following ABX exposure. **A.** Schematic representation of experimental plan. **B.** Alpha diversity analysis of stool microbiome diversity in mice (n=5/group). **C.** UniFrac analysis depicting the clustering of bacterial communities studied. Each point represents a community from a single mouse. **D.** Relative microbial composition by family through dietary and antibiotic interventions.

Beta diversity analyses of community composition demonstrated that the baseline fecal microbiota was similar across groups prior to diet or antibiotic interventions. Unifrac analysis revealed ABX treated animals clustered separately from animals that did not receive AVNM. During the recovery phase, PBEN and AEN mice exhibited distinct clustering patterns compared to the antibiotic naïve and antibiotic treated groups (Figure 1C). Notably, the gut microbiota of PBEN-fed mice recovering from antibiotics showed greater similarity to ABX-naive animals, while the AEN-fed mice exhibited profoundly deranged microbial communities even during the recovery phase. These results show how the gut microbial community is significantly altered by antibiotics and can be shaped by diet during ABX recovery. Specifically, PBEN supports microbiota richness while AEN leads to loss of diversity independently and after perturbations with ABX.

We also observed diet-dependent taxonomic differences in gut microbial profiles (Figure 1D). In the absence of antibiotics, PBEN fed animals harbored more short chain fatty acid producing commensals including Bacteroidetes, Ruminococcaceae, and Lachnospiraceae. AEN samples were enriched with bacteria with pathogenic or mucus-degrading potential such as Erysipelotrichaceae^24^ and Verrucomicrobiaceae^25,26^, which have been shown previously to bloom with fiber deficient diets^27^. After one week of AVNM treatment, PBEN and AEN communities were both markedly disrupted but PBEN randomized mice showed more favorable recovery with restoration of beneficial commensals whereas AEN randomized samples were almost uniformly dominated by Enterobacteriaceae. Confirmation experiments with a different cohort of animals also revealed the presence of pathogens in post-antibiotic AEN samples (Enterobacteriaceae and Enterococcaceae) (not shown). These results suggest PBEN is superior to AEN in mitigating ABX-induced gut dysbiosis.

Two additional iterations of this experiment were conducted, varying the diets during the antibiotic and recovery phases. Here, either all animals received AEN instead of normal chow during the initial week of antibiotics (Figure S1A), or half received PBEN and half received AEN during antibiotic treatment (Figure S1B). In both cases, PBEN was consistently superior in maintaining alpha diversity and mitigating dysbiosis compared to AEN. Alpha diversity reproducibly collapsed with antibiotic exposure, but the collapse was less severe and less prolonged with PBEN relative to AEN. Even without antibiotic exposure, alpha diversity decreased significantly with AEN and was maintained with PBEN. Thus, as shown in our prior studies, AEN promotes dysbiosis relative to PBEN, and these differences are accentuated in the presence of ABX.

### AEN sustains colitis during recovery from ABX treatment and promotes bacterial dissemination

Gastrointestinal distress is a well-documented and at times debilitating consequence of antibiotic treatment in both rodents and humans^3,28^. Animals receiving antibiotics exhibited signs of intestinal distress similar to those commonly seen in colitis models. The severity of colitis was assessed using a modified disease activity index (DAI) by scoring weight loss, stool consistency, and hematochezia. Regardless of diet, animals exposed to AVNM uniformly had increased DAI scores (Figure 2A). Animals receiving PBEN during the washout period reliably experienced an improvement in symptoms whereas disease severity persisted among animals receiving AEN. Cecal size and weight^28^ increased markedly with antibiotic treatment and this increase was far greater in AEN fed animals (Figure 2B). As also reported by others^29^, histological examination of colonic tissue revealed no overt intestinal damage after ABX (not shown). However, we observed enlarged submucosal lymphoid aggregates after gut recolonization, especially in AEN fed mice (Figure S2A). Such lymphoid structures have been associated with inflammatory bowel disease are strongly suggestive of a gut-specific immune response^30–32^. Finally, we measured fecal lipocalin (LCN2) as a biomarker of intestinal inflammation^33^. As previously shown^34,35^, ABX treatment decreased LCN2 levels. Interestingly, repopulation of the gut during the washout period led to a marked increase in LCN2 in all groups, but the rise was more pronounced in AEN-fed animals (Figure 2C). These results indicate that, by some measures, the washout period after ABX cessation is associated with more gut inflammation than the ABX treatment period. Importantly, this post-ABX inflammation is diet dependent.

**Figure 2.**
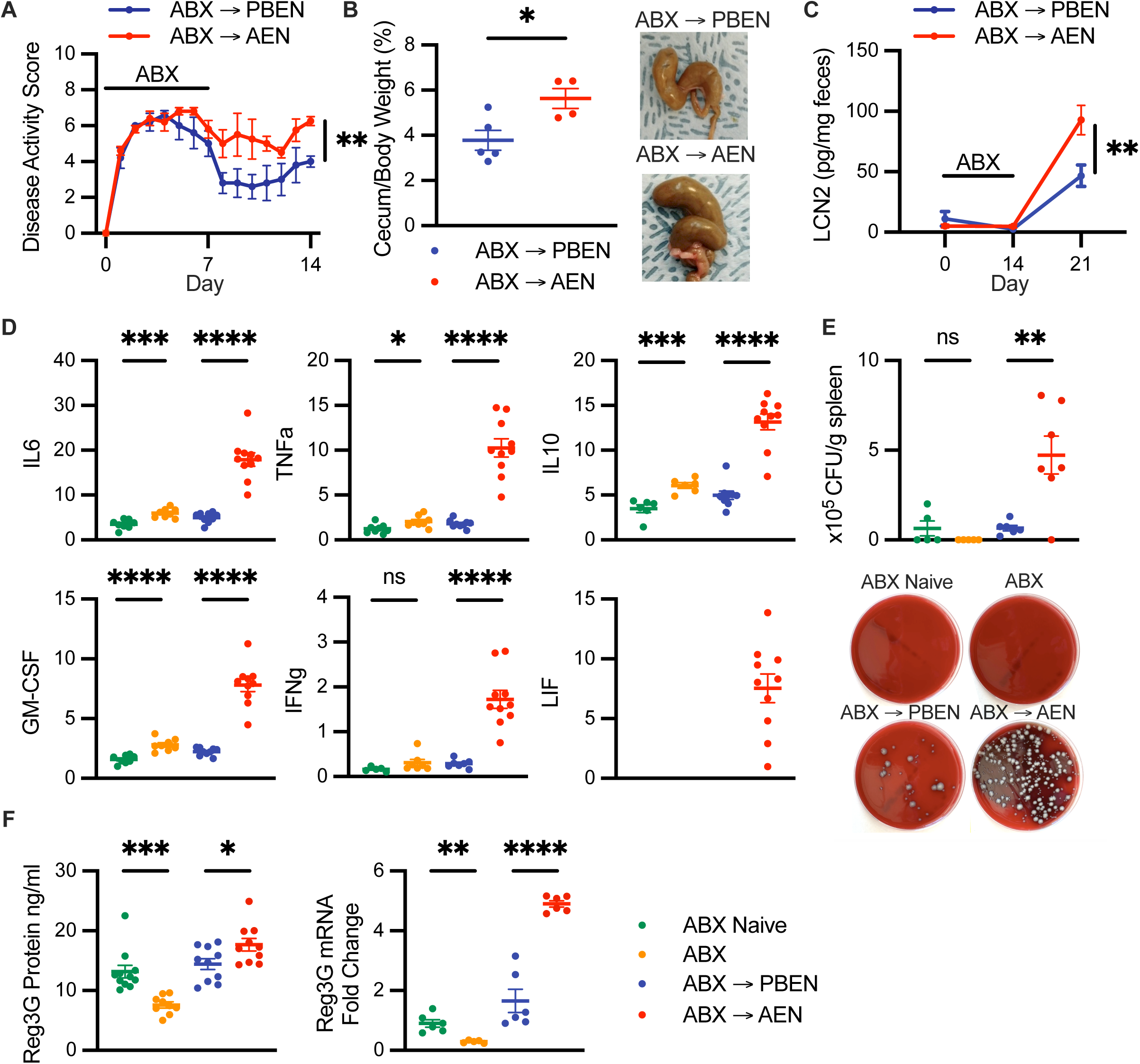
PBEN but not AEN mitigates signs of intestinal inflammation due to ABX exposure. A. Disease activity index (DAI) scores in animals undergoing antibiotic (ABX) treatment followed by randomization to PBEN or AEN (n=5/group). B. Cecal to body weight ratio in animals receiving PBEN (n=5) or AEN (n=4) diet during ABX treatment with representative images of ceca from different groups. C. Concentration of LCN2 in feces from mice in the treatment groups (n=11/group). D. Colon wall cytokine levels including IL6, TNFα, IL10, GM-CSF, IFNγ, LIF in experimental groups shown as pg/mg of protein (n=6-11/group). E. Bacterial burden in the spleen (n=5-7/group) with representative images. F. Ileal Reg3G protein quantified by ELISA and mRNA through quantitative RT-PCR in experimental groups (n= 5-11/ group). Data represent mean ± standard error of the mean (SEM) with points representing individual mice. Statistical significance determined by Student t test between ABX naive versus ABX treated and PBEN versus AEN after antibiotic treatment from at least two independent experiments. ns, not significant; \**P* < .05, \*\**P* < .01, \*\*\**P* < .001, \*\*\*\**P* < .0001.

To better understand ABX-induced intestinal inflammation and diet dependent recovery, we studied changes in cytokine production in the colon wall (Figure 2D). In some cases, e.g. IL6 and TNF-alpha, we observed modest antibiotic-induced increases in pro-inflammatory cytokine concentration. However, we observed much higher increases during the post-antibiotic washout period in cytokines including G-CSF, GM-CSF, IFNg, TNFa, IL10, IL6, and these surrogates for inflammation were particularly increased in samples receiving AEN. Interestingly, LIF (leukemia inhibitory factor), a member of the IL-6 superfamily secreted by inflamed gut epithelial cells in response to dysbiosis^36,37^, was undetectable in all samples except AEN samples collected during the washout period. To test the hypothesis that reestablishment of the gut microbiome contributes to the post-antibiotic inflammatory response, we developed a co-culture system in which immortalized colonic epithelial cells were incubated with fecal samples from mice treated with antibiotics, followed by either PBEN or AEN during the washout period and analyzed expression of IL8, a mediator of GI inflammatory response^38–40^. After one week of recovery, both diets elicit expression of IL8, suggesting recolonization to be a pro-inflammatory event. IL8 expression was higher with AEN samples than PBEN, although this did not reach statistical significance. Coculturing HT29 with heat-killed bacteria from the same stool reduced this difference (Figure S2B), indicating that viable bacteria within fecal samples are responsible for the observed diet-dependent effects.

Gut inflammation weakens the intestinal barrier, allowing dissemination of bacteria and their byproducts in experimental animals and patients^41,42^. Similarly, antibiotic-induced microbiota depletion also compromises gut integrity and permits bacterial translocation^43–47^. We therefore evaluated bacterial colony-forming units (CFUs) recovered within the spleens of antibiotic-treated animals before and after randomization to the experimental diets. As expected, only minimal bacterial CFUs were recovered from spleens of ABX naïve or ABX treated animals. However, we observed a substantial growth of bacteria within spleen samples collected 1 week following cessation of ABX (Figure 2E). Animals fed AEN during the antibiotic washout period exhibited heavy bacterial burdens in the spleen, far surpassing the levels observed in controls or in animals randomized to PBEN. These data confirm prior reports that bacterial translocation from the gut peaks *after* cessation of ABX^43,47^ and also indicate that dietary intervention can modulate the magnitude of bacterial translocation after ABX. Given the known role of *MUC2* and *Reg3G* as regulators of mucosal immunity and intestinal inflammation, we hypothesized that their expression will be altered in response to ABX-induced changes in the gut microbiome. In line with observations by other groups that ABX disrupts the colonic mucus barrier^48,49^, we found ABX treatment decreases colonic MUC2 expression that remained low even after the one week recovery period in both diet groups (Figure S2C). Similarly, we confirmed previous findings that ABX treatment decreases ileal Reg3 expression^50^ (Figure 2F). As with fecal lipocalin, increase in Reg3g was far more significant with AEN compared to PBEN, likely in response to particular microbes blooming during the recolonization phase given the bactericidal activity of Reg3.

### PBEN is superior to AEN in rescuing ABX-induced anemia and leukopenia

In the absence of ABX perturbations, animals randomized to receive PBEN and AEN have similar complete blood count (CBC) profiles (Figure S3A). As reported by others^51^, we also found that antibiotic induced dysbiosis disrupts hematopoiesis leading to anemia and leukopenia (Figure 3B). However, mice fed PBEN during the recovery phase exhibited significant improvements in anemia, evident by increased hemoglobin levels compared to those fed AEN who remained anemic. ABX treatment also resulted in neutrophilia and leukopenia, resulting in a higher neutrophil-to-lymphocyte ratio (NLR), a marker associated with increased mortality in intensive care units^52–55^. While lymphocyte counts during the post-ABX washout phase were comparable between PBEN and AEN fed mice, AEN mice displayed significantly higher neutrophil counts resulting in elevated NLR.

**Figure 3.**
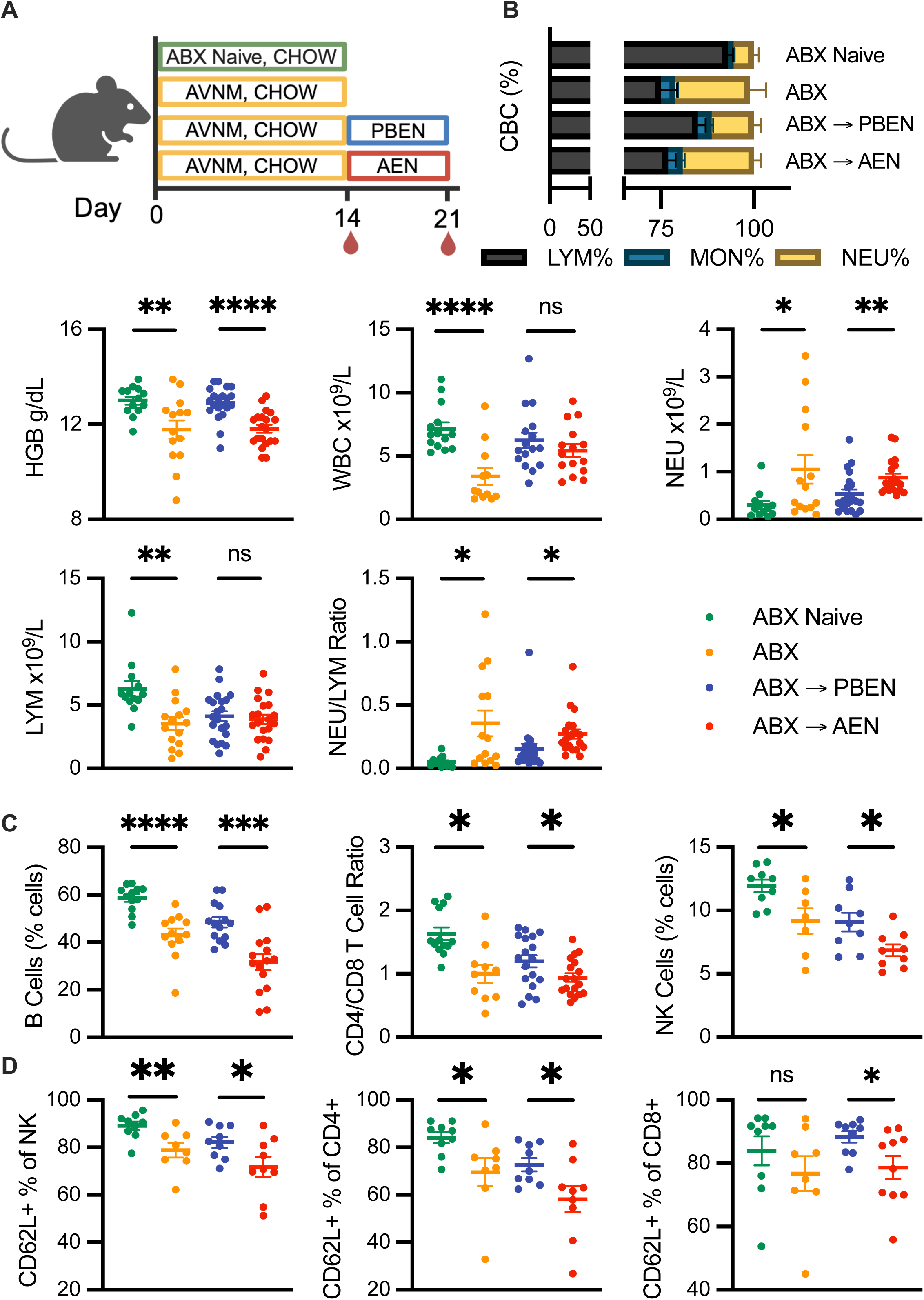
PBEN is superior to AEN in rescuing ABX-induced anemia and leukopenia. A. Schematic of experimental plan. B. Peripheral blood cell profiles following antibiotic (ABX) treatment and dietary interventions including complete blood count differential, hemoglobin concentration, white blood cell (WBC), neutrophil (NEU) and lymphocyte (LYM) counts as along with the neutrophil to lymphocyte ratio. C. Flow cytometric analysis of lymphocyte subsets in peripheral blood following antibiotic (ABX) treatment and dietary intervention showing B cells, CD4/CD8 ratio, and natural killer (NK) cells. Proportion of CD62L+ NK cells, CD4 and CD8 T cells signifying naïve lymphocytes also shown. Data represent mean ± standard error of the mean (SEM) with points representing individual mice. Statistical significance was determined by Student t test between ABX naive versus ABX treated and PBEN versus AEN after antibiotic treatment from at least two independent experiments (n=8-15/ group). ns, not significant; \**P* < .05, \*\**P* < .01, \*\*\**P* < .001, \*\*\*\**P* < .0001.

Using flow cytometry, we found that ABX treatment led to a decrease in B (CD11b-B220+) cells and natural killer (NK) (CD11b-NK1.1+) cells. As reported by others^51^, CD4 (CD11b-CD4+) T cells also decreased but CD8 (CD11b-CD8a+) T cells increased, resulting in a lower CD4/CD8 ratio (Figure 3C). During recovery, PBEN was superior in mitigating most antibiotic induced derangements and restoring lymphoid cells. AEN-fed mice displayed further decline in B and NK cell levels during the washout, along with a lower CD4/CD8 ratio compared to ABX-treated mice. We further investigated the activation state of these lymphocytes using CD62L (L-selectin) expression, a marker highly expressed on naive cells which is shed in the process of homing to sites of inflammation^56–58^. ABX treatment decreased the proportion of naive NK, CD4+, and CD8+ T cells, indicated by a decrease in CD62L+ population (Figure 3D). During recovery, PBEN partially restored these naive populations, whereas AEN-fed mice exhibited a continued decrease in naïve NK and CD4+ T cells or remained low in CD8+ T cells. These findings collectively suggest that ABX treatment significantly disrupts peripheral immunity, favoring granulocyte over lymphocyte populations and deranging the balance between naive and effector lymphocytes. PBEN diet appears to be more effective than the AEN diet in mitigating these effects and promoting hematologic recovery.

### AEN sustains an expansion of myeloid cells and contraction of lymphoid cells in the bone marrow after ABX

To determine if observed changes in peripheral blood reflect alterations in bone marrow hematopoiesis, we assessed bone marrow cellularity and performed flow cytometric analysis of mature cells. Consistent with previous studies^51^, ABX treatment drastically reduced bone marrow cellularity. PBEN fed mice displayed higher bone marrow cellularity compared to AEN during the recovery phase after ABX cessation (Figure 4A). Pan-myeloid marker CD11b staining revealed an overall increase in myeloid cell frequency within the bone marrow after ABX treatment. Importantly, PBEN more effectively mitigated this increase during recovery compared to AEN (Figure 4B). Antibiotic treatment also significantly reduced bone marrow B cells and the CD4/CD8 ratio as previously reported^51^. During recovery, PBEN mice exhibited improved B cell restoration (Figure 4C). Although neither diet facilitated complete recovery of CD4 or CD8 populations, the CD4/CD8 ratio was normalized in PBEN fed mice but increased significantly in the AEN group (Figure 4D). These findings indicate that ABX results in significant alterations in mature bone marrow cells present that are partially resolved with PBEN diet but exacerbated with AEN.

**Figure 4.**
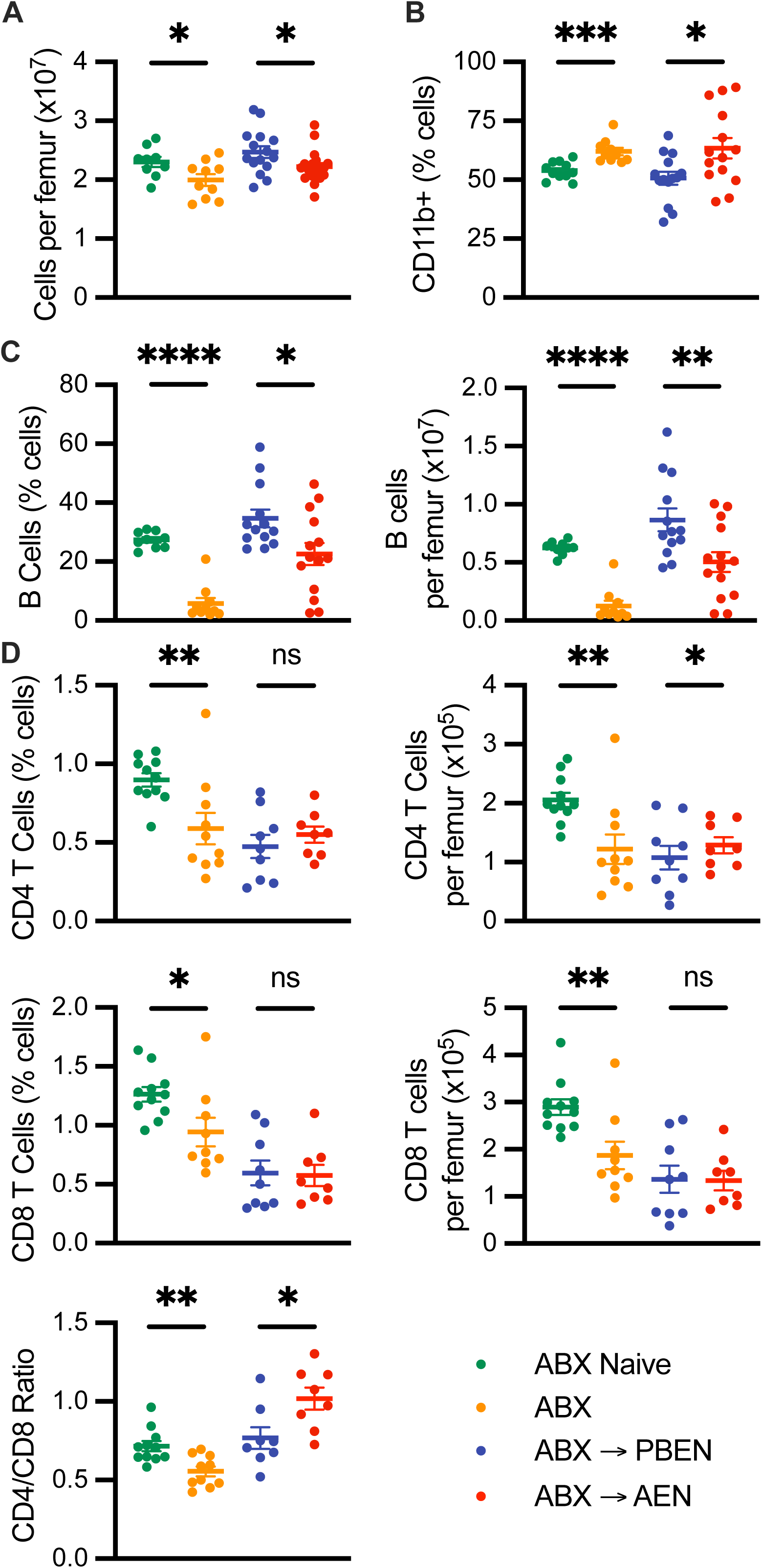
AEN sustains bone marrow shift towards myeloid at the expense of lymphoid cells post ABX. A. Bone marrow (BM) cellularity following antibiotic (ABX) treatment and after ABX with dietary interventions. B. Pan-myeloid cell frequency (CD11b+) C. B cells frequencies and count and D. CD4 and CD8 T cell frequencies and total counts per femur with bone marrow CD4/CD8 ratio. All data represent mean ± standard error of the mean (SEM) with points representing individual mice. Statistical significance was determined by Student t test between ABX naive versus ABX treated and PBEN versus AEN after antibiotic treatment from at least two independent experiments (n=7-14/ group). ns, not significant; \**P* < .05, \*\**P* < .01, \*\*\**P* < .001, \*\*\*\**P* < .0001.

### PBEN minimizes severity of post-antibiotic vancomycin-resistant *Enterococcus faecalis* and *Klebsiella pneumoniae* GI infections but not *Klebsiella* induced pneumonia

After observing ABX and diet induced alterations in white and red blood cells in peripheral blood and bone marrow, we investigated whether diet choice impacts susceptibility to infectious and inflammatory challenges relevant to hospitalized patients receiving ABX and enteral nutrition. Using a previously established model of AID and breakdown of the gut barrier^59^ we tested the impact of PBEN and AEN on bacterial clearance and systemic translocation. PBEN and AEN fed mice were treated with neomycin and vancomycin for three days then inoculated with *K. pneumoniae* 396 (KP) by gastric gavage. Intestinal inflammation was provoked 14 days post infection (dpi) using dextran sodium sulfate (DSS) to drinking water for three days. Fecal pellets were collected throughout the experiment to monitor KP colonization and clearance patterns, and animals were sacrificed 3 days post DSS to assess KP dissemination to the spleen. Fecal KP burdens were similar in PBEN and AEN mice at 3 dpi. At sacrifice on 15 dpi, fecal KP burden, fecal lipocalin, and bacterial translocation to the spleen were significantly higher in AEN fed mice relative to PBEN fed mice (Figure 5A and Figure S4A).

**Figure 5.**
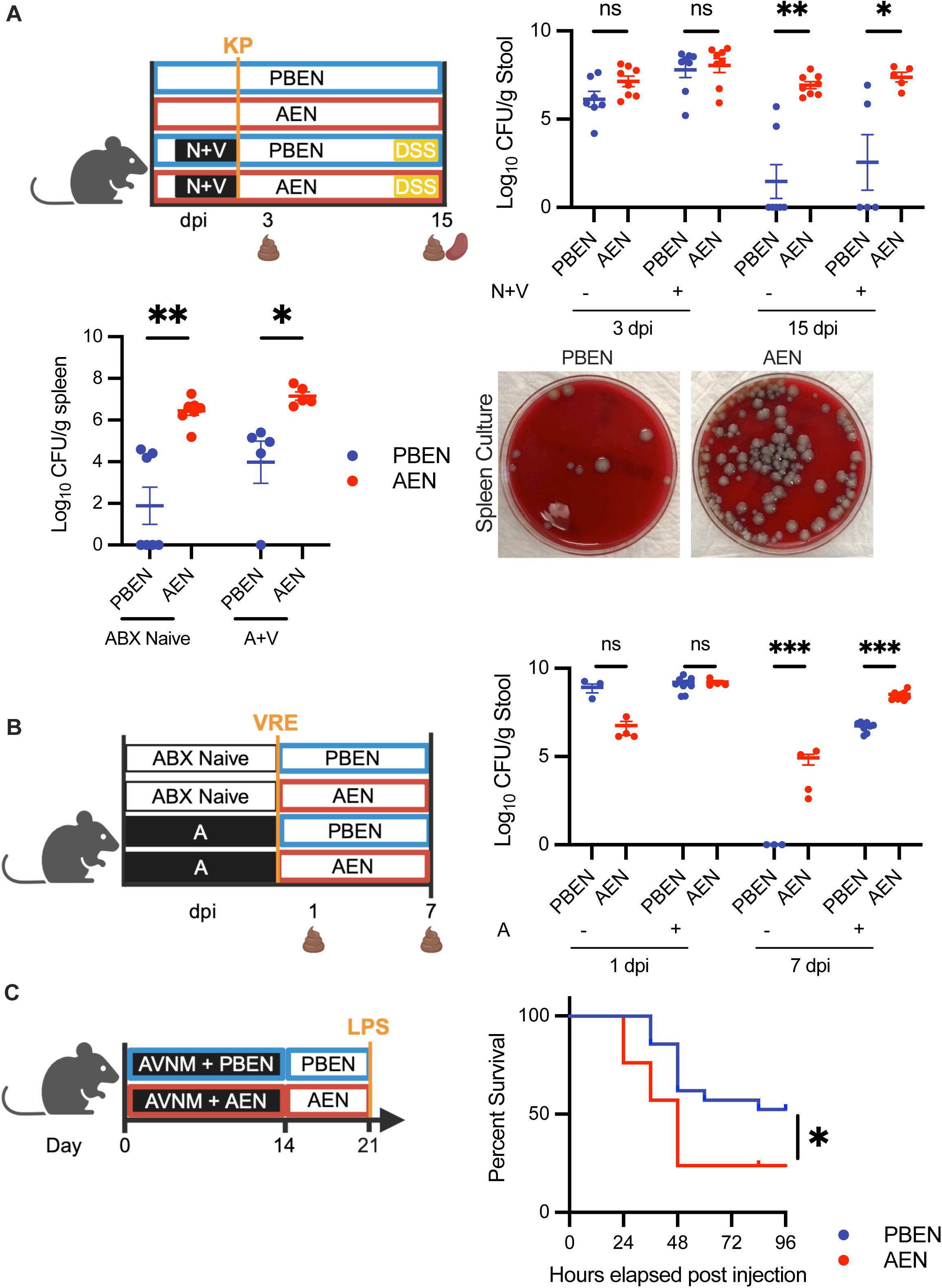
PBEN minimizes severity of post-antibiotic clearance of vancomycin-resistant *Enterococcus faecalis* and *Klebsiella Pneumoniae* GI infections but not *Klebsiella* induced pneumonia. A. Schematic of *Klebsiella Pneumoniae-*DSS experiment. qPCR was used to quantify KP396 in stool 3 and 5 dpi (n=5-8/group). KP396 was also quantified in the spleen (n=5/group). Representative photos of spleen cultures showing bacterial dissemination also shown. B. Schematic of Vancomycin-Resistant *Enterococcus faecalis* (VRE) experimental plan and cecal VRE burden was assessed on 1 and 7 dpi (n=4-9/group) C. Schematic of lipopolysaccharide (LPS) experiment. After ABX and diet interventions, animals were subjected to a lethal LPS injection and followed for 96 hours (n=21/diet). Kaplan-Meier survival curve comparing survival of PBEN and AEN fed mice following LPS challenge (n=4 mice/diet). Survival analyzed using the Log-rank (Mantel-Cox) statistical test. All other data represent mean ± standard error of the mean (SEM) with points representing individual mice. Student t test between PBEN versus AEN randomized. ns, not significant; \**P* < .05, \*\**P* < .01, \*\*\**P* < .001, \*\*\*\**P* < .0001.

Interestingly, the protection offered by PBEN was not conferred in a KP induced pneumonia model where after two weeks of ABX and one week of washout with either PBEN or AEN, mice were subjected to intratracheal KP inoculation. In this model, AEN fed mice had more favorable outcomes than PBEN (Figure S4B) suggesting protection might be limited to GI infections. Therefore, we recapitulated another published model to determine whether the observed protective benefits of PBEN against gram-negative KP also extended to another common nosocomial pathogen in the gut. Mice were randomized into four groups receiving either PBEN or AEN with or without ampicillin for seven days. Upon cessation of ABX, animals continued on either PBEN or AEN during a 7 day washout period. At the conclusion of the washout period, all groups were gavaged with 10^5^ CFUs of Vancomycin-Resistant *Enterococcus faecalis* (VRE)^60^. While both PBEN and AEN groups treated with antibiotics showed no VRE before infection and similar levels at 1 dpi, PBEN-fed mice displayed significantly lower VRE burden 7 dpi compared to AEN (Figure 5B). Interestingly, even in the absence of antibiotics, PBEN fed mice completely cleared the infection, whereas AEN-fed mice continued to harbor VRE. These results suggest that PBEN offers a distinct advantage over AEN in promoting clearance of intestinal VRE.

In a third evaluation of diet-dependent responses to a post-ABX inflammatory stimulus, mice received AVNM for two weeks followed by one week recovery on PBEN or AEN. On the final day of recovery, they received an IP injection of a potentially lethal dose of lipopolysaccharide (LPS) and were then monitored for survival. Over the course of 3 separate cohorts, PBEN fed mice demonstrated reproducibly increased survival compared to AEN (Figure 5C). PBEN resulted in 37.5% mortality with survivors remaining healthy, while AEN resulted in 62.5% mortality with a median survival time of only 48 hours. These findings suggest PBEN offers a more robust response to inflammatory challenges and a protective effect against LPS-induced lethality. In another iteration of this experiment, we found higher plasma IL-10 and lower TNFa in PBEN mice compared to AEN two hours post LPS injection (Figure S4C). Overall, our data suggest that PBEN after antibiotic treatment offers a distinct advantage in promoting response and recovery of a robust inflammatory response and bacterial clearance in some but not all models.

### Dietary intervention with pediatric PBEN promotes microbiome recovery and improved peripheral blood profile in critically ill children

We previously demonstrated the severity of ICU dysbiosis in critically ill human patients, and separately we have also shown that open-label conversion from AEN to PBEN in pediatric outpatients promotes the growth of commensal gut anaerobes and decreases the prevalence of pathogen colonization in the gut^18,61–64^. Here, we conducted a randomized trial of children admitted to a single quaternary care pediatric intensive care unit (PICU). All subjects were patients initiated on enteral feeds via a feeding tube by the treating physician as part of routine clinical practice with no other changes to medical management. After obtaining written consent, children were randomized in a 1:1 ratio to commercially available pediatric PBEN (Nourish®, Functional Formularies, West Chester, OH, USA) or AEN (Pediasure®, Abbott Nutritional Products, Abbott Park, IL, USA).

Thirty-two patients were recruited and randomized with 14 patients randomized to 2 weeks of Pediasure and 18 patients randomized to 2 weeks of Nourish. Of these, 28 patients completed the study with 13 patients on the Pediasure diet and 15 on Nourish. The time elapsed between admission and initiation of enteral feeds was determined by the clinical team without input from the research study team. Baseline characteristics for the 28 patients who completed the study including race, sex, body mass index, percentage of patients with co-morbid condition, and PIM-3 score were similar (**Table 1**). Children randomized to Pediasure were younger (3.1y ± 2.3 vs. 6.1y ± 4.2; p=0.03). Clinically, there was no difference between treatment groups in antibiotic exposure, positive cultures, or percentage of patients receiving parenteral nutrition.

**Table 1.**
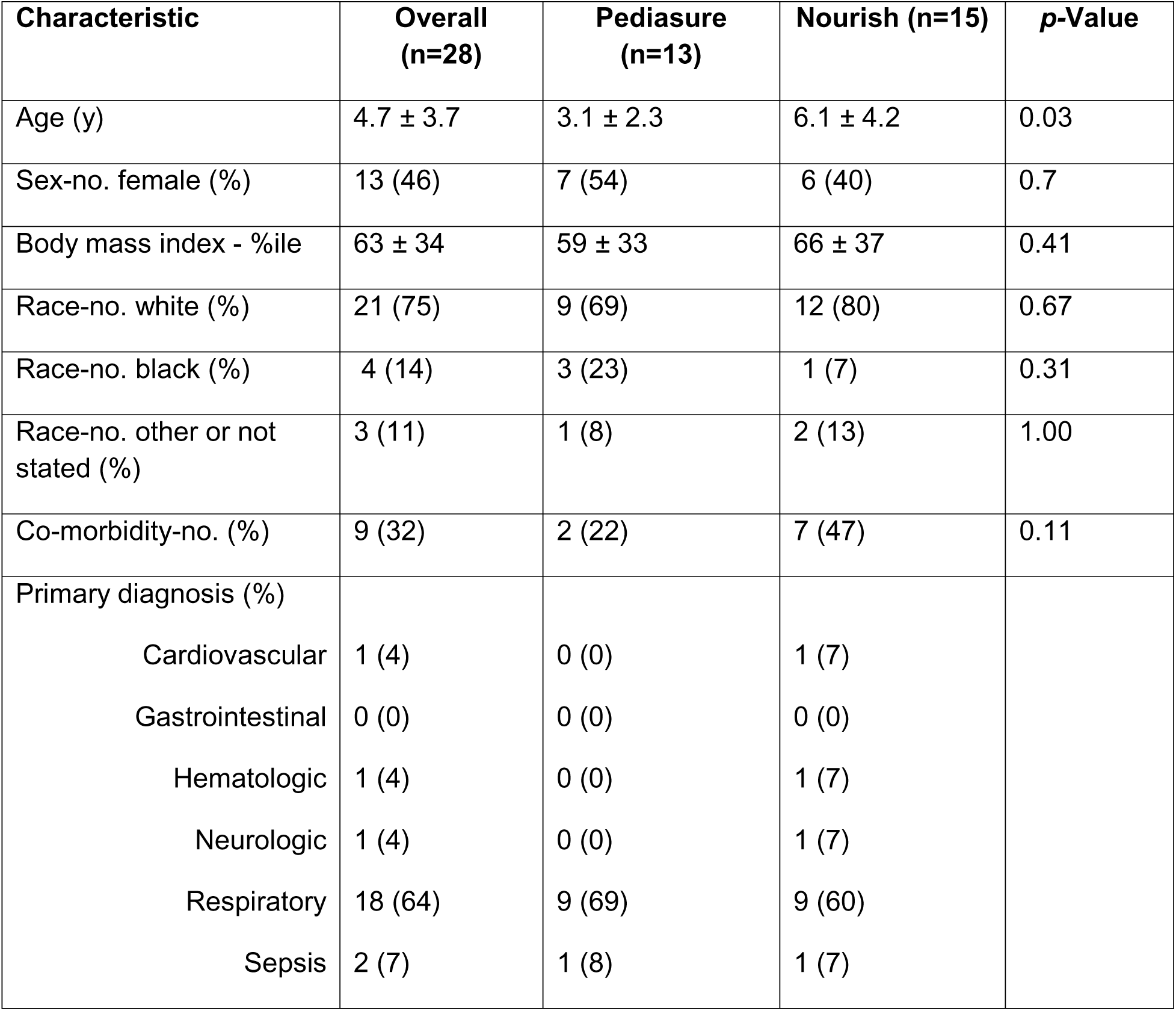
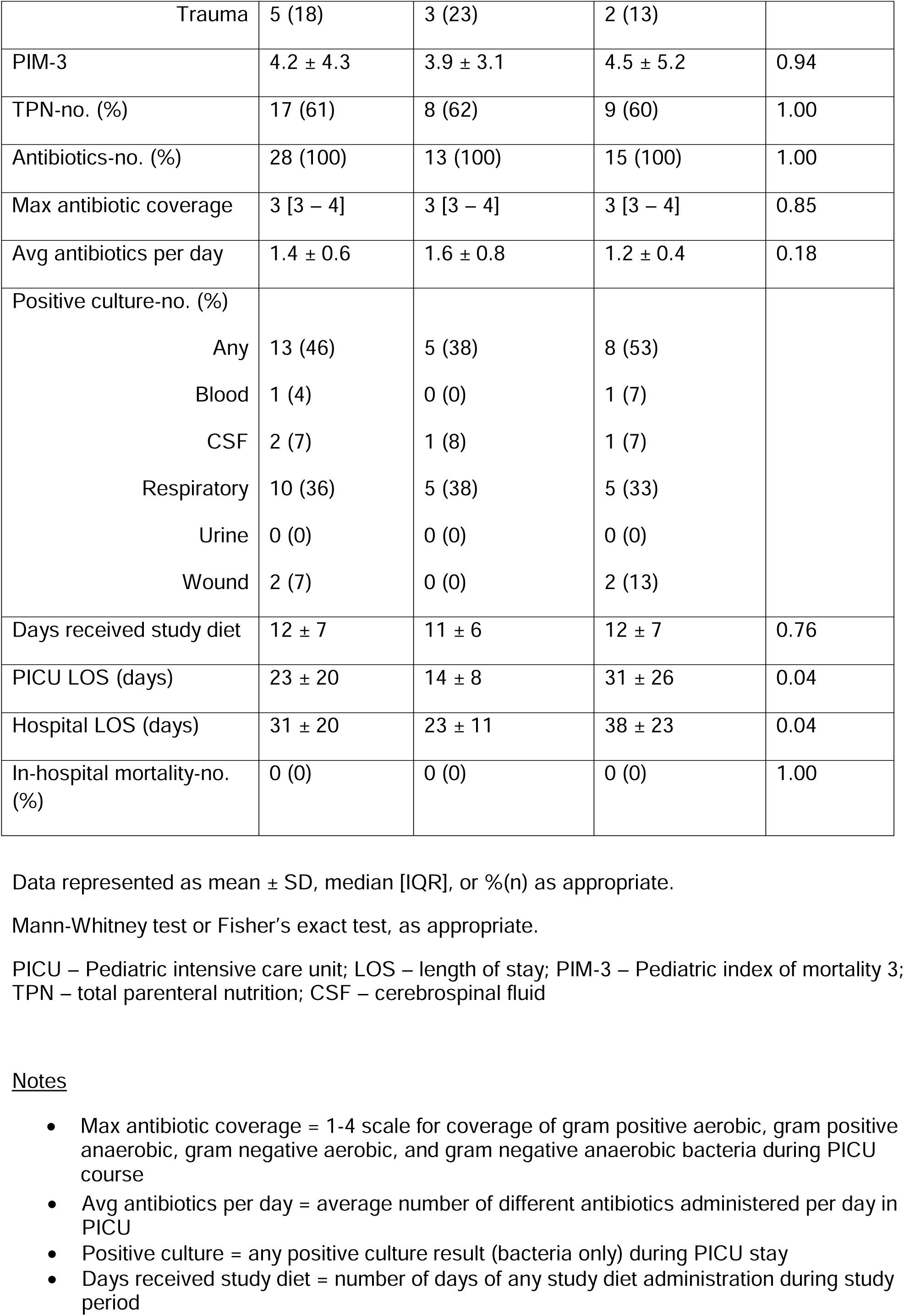
Characteristics of patients in clinical trial.

| Characteristic | Overall (n=28) | Pediasure (n=13) | Nourish (n=15) | p-Value |
| --- | --- | --- | --- | --- |
| Age (y) | 4.7 ± 3.7 | 3.1 ± 2.3 | 6.1 ± 4.2 | 0.03 |
| Sex-no. female (%) | 13 (46) | 7 (54) | 6 (40) | 0.7 |
| Body mass index - %ile | 63 ± 34 | 59 ± 33 | 66 ± 37 | 0.41 |
| Race-no. white (%) | 21 (75) | 9 (69) | 12 (80) | 0.67 |
| Race-no. black (%) | 4 (14) | 3 (23) | 1 (7) | 0.31 |
| Race-no. other or not stated (%) | 3 (11) | 1 (8) | 2 (13) | 1.00 |
| Co-morbidity-no. (%) | 9 (32) | 2 (22) | 7 (47) | 0.11 |
| Primary diagnosis (%) |  |  |  |  |
| Cardiovascular | 1 (4) | 0 (0) | 1 (7) |  |
| Gastrointestinal | 0 (0) | 0 (0) | 0 (0) |  |
| Hematologic | 1 (4) | 0 (0) | 1 (7) |  |
| Neurologic | 1 (4) | 0 (0) | 1 (7) |  |
| Respiratory | 18 (64) | 9 (69) | 9 (60) |  |
| Sepsis | 2 (7) | 1 (8) | 1 (7) |  |
| Trauma | 5 (18) | 3 (23) | 2 (13) |  |
| PIM-3 | 4.2 ± 4.3 | 3.9 ± 3.1 | 4.5 ± 5.2 | 0.94 |
| TPN-no. (%) | 17 (61) | 8 (62) | 9 (60) | 1.00 |
| Antibiotics-no. (%) | 28 (100) | 13 (100) | 15 (100) | 1.00 |
| Max antibiotic coverage | 3 [3 – 4] | 3 [3 – 4] | 3 [3 – 4] | 0.85 |
| Avg antibiotics per day | 1.4 ± 0.6 | 1.6 ± 0.8 | 1.2 ± 0.4 | 0.18 |
| Positive culture-no. (%) |  |  |  |  |
| Any | 13 (46) | 5 (38) | 8 (53) |  |
| Blood | 1 (4) | 0 (0) | 1 (7) |  |
| CSF | 2 (7) | 1 (8) | 1 (7) |  |
| Respiratory | 10 (36) | 5 (38) | 5 (33) |  |
| Urine | 0 (0) | 0 (0) | 0 (0) |  |
| Wound | 2 (7) | 0 (0) | 2 (13) |  |
| Days received study diet | 12 ± 7 | 11 ± 6 | 12 ± 7 | 0.76 |
| PICU LOS (days) | 23 ± 20 | 14 ± 8 | 31 ± 26 | 0.04 |
| Hospital LOS (days) | 31 ± 20 | 23 ± 11 | 38 ± 23 | 0.04 |
| In-hospital mortality-no. (%) | 0 (0) | 0 (0) | 0 (0) | 1.00 |

Data represented as mean ± SD, median [IQR], or %(n) as appropriate.
Mann-Whitney test or Fisher's exact test, as appropriate.
PICU – Pediatric intensive care unit; LOS – length of stay; PIM-3 – Pediatric index of mortality 3; TPN – total parenteral nutrition; CSF – cerebrospinal fluid

Metagenomic sequencing of microbial DNA from fecal samples and rectal swabs confirmed that baseline microbial diversity was similar across groups. By contrast, at the conclusion of feeding, microbiome profiles differed markedly between groups (Figure S5D). The most abundant taxa across the entire cohort of PBEN samples included *Bacteroides ovatus* and *Phocaeicola vulgatus* (previously *B. vulgatus),* established commensal anaerobes that degrade plant fiber to generate metabolites such as N-methylserotonin and short chain fatty acids associated with protective intestinal inflammatory responses^65–67^. By contrast, the most abundant taxa within samples from patients receiving AEN were the potential pathogens *Candida albicans* and *Streptococcus* (Figure 6A). SCFA producers from the phylum Bacteroidetes including *Bacteroides ovatus*, *Phocaeicola vulgatus, and Phocaeicola dorei* were far less prevalent in AEN fecal samples. To characterize the functional properties of the respective microbial populations, we used the Carbohydrate-Active enZymes database (CAZy) to identify diet-dependent enzymes involved in carbohydrate metabolism. Overall, CAZy coding genes were highly upregulated (ie higher total gene counts) in PBEN patients compared to AEN (Figure 6B). Highly enriched CAZy genes in PBEN samples included polygalacturonase, a pectin degrading enzyme^68^, and beta glucosidase, which degrades dietary fiber in the final step of hydrolysis by converting cellobiose to glucose^69^ (Figure 6C). Our findings demonstrate that dietary intervention with PBEN in critically ill children can promote microbiome recovery, reduce the abundance of potential pathogens, and enhance the growth of beneficial commensal bacteria, ultimately leading to improved peripheral blood profiles and potentially better clinical outcomes.

**Figure 6.**
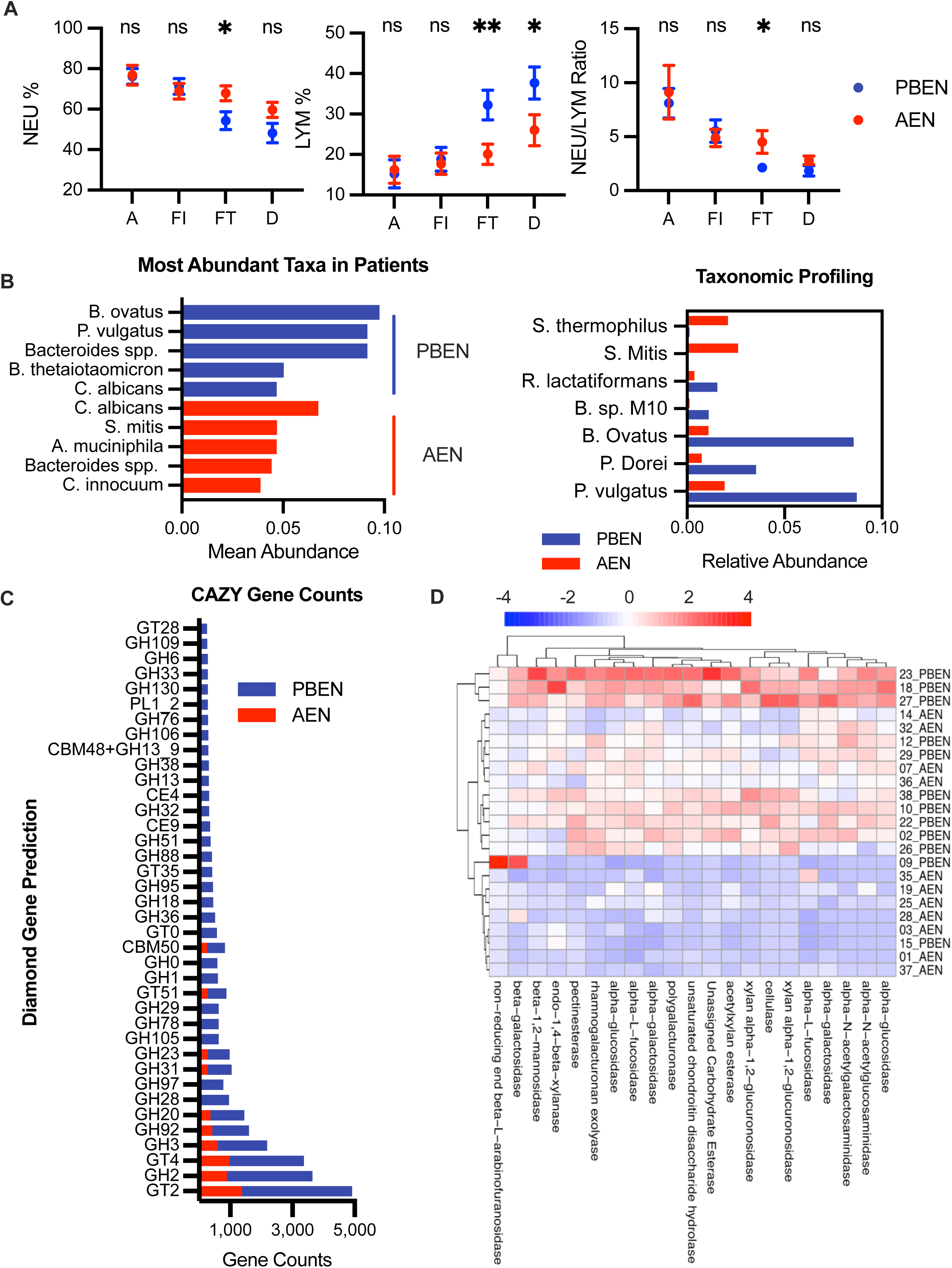
Dietary intervention with pediatric PBEN promotes microbiome recovery and improved peripheral blood profile in randomized clinical trial in critically ill children. A. Most abundant taxa and B. relative abundance of microbes in in patients randomized to PBEN or AEN. C. CAZy coding genes enriched in both diets with gene counts >250 E. Polygalacturonase and beta glucosidase enrichment in PBEN samples compared to AEN. D. Percentage of white blood cells that are neutrophils (NEU) and lymphocytes (LYM) along with neutrophil to lymphocyte ratio at time of admission, feed initiation, feed termination, and hospital discharge.

**Figure 7.**
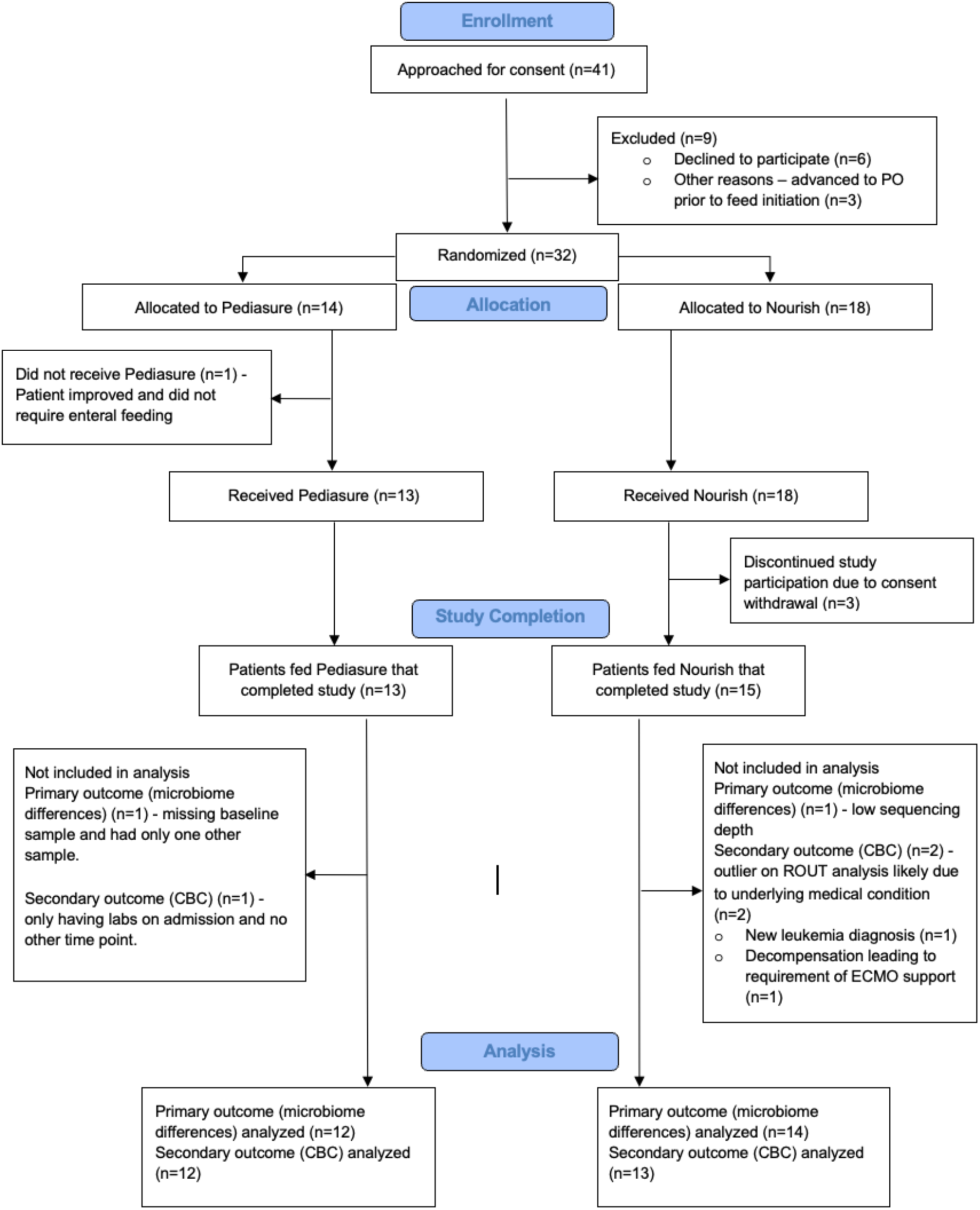
Modified CONSORT diagram. Flowchart of participants disposition throughout the study.

Importantly, all enrolled subjects received antibiotics, and there were no appreciable differences in antibiotic exposure between the PBEN and AEN groups. Similarly, there were no significant differences in white blood cell (WBC) count, platelet count, or CRP and white blood cells (WBC) at admission (Figure S5B). There were no differences in lymphocytes or neutrophils at admission or study diet initiation between groups. After the 2 week diet intervention, patients in the PBEN group had a higher percentage of WBC that were lymphocytes and lower percentage of WBC that were neutrophils (Figure 6D). Collectively, these results suggest diet can influence microbiome composition and peripheral blood immune populations in critically ill patients.

## Discussion

Nutritional supplementation for hospitalized patients that cannot eat by mouth remains controversial due to concerns about optimal timing, route, and quality of supplementation. Over the past century, enteral formulas with chemically defined artificial compositions have replaced older practices of administering pureed food to patients, providing precise calorie and macronutrient information. However, factors such as ingredient sourcing and quality have been largely overlooked, leaving critical aspects of nutrition unaddressed. No studies have investigated how different types of nutrition in hospitalized patients affect the microbiome or clinical outcomes.

Natural, whole foods-based alternatives to conventional enteral nutrition have emerged, offering putative benefits of higher fiber content, lower sugar content, and the complete absence of artificial emulsifiers and preservatives. These formulas may be less likely to induce dysbiosis and gut inflammation known to result from dietary exposure to the added sugars, artificial sweeteners, and chemical emulsifiers present in AEN^17,70,71^. In theory, formulas with high fiber content should provide valuable substrates for growth of beneficial short chain fatty acid-producing gut bacteria and promote a healthy gut microbiome. Despite these potential advantages as well as microbiome-independent benefits of fiber intake^72^, the impact of these natural plant-based formulas in clinical medicine has not been extensively studied.

This study aimed to compare how commercially available PBEN and AEN affect the detrimental impact of antibiotics. This is an understudied topic of high clinical importance as a significant percentage of ICU patients are exposed to antibiotics and fed with AEN, and it is plausible that nutritional strategies to simultaneously support both host and microbe could improve ICU outcomes. A precedent for such an idea is provided by recent studies demonstrating that dietary fiber improves outcomes^14^ and antibiotics worsens outcomes among patients undergoing treatment for cancer ^73,74^.

Using a sequencing-based approach, we found that gut microbiome profiles between animals eating PBEN and AEN diverge significantly, even without antibiotics. The PBEN microbiome is more diverse and contains a higher abundance of commensal anaerobes such as Lachnospiraceae. Prior studies in humans with Crohn’s Disease have consistently demonstrated decreased diversity with AEN, albeit with conflicting results regarding gut pathogen abundance^75^. Diet-dependent differences were accentuated with antibiotics. These results align with previous studies showing that high-fat or fiber deficient diets exacerbates AID through various mechanisms^76–79^, while a high-fiber diet can mitigate it^35,80^.

Prior studies have documented ABX-induced gut barrier breakdown^32,46^ and increased susceptibility to colitis; in fact, some groups intentionally use ABX to induce gut inflammation in rodent models of IBD^47^. Our observations of gross evidence of intestinal distress across cohorts of ABX-treated mice validate these findings. We further showed that gut and systemic inflammation worsen during the recolonization phase after stopping ABX, as evidenced by increased fecal lipocalin, cytokine production in the colon, and translocation of bacteria to the spleen. This is consistent with reports by Knoop et al^47^, who also found bacterial translocation from the gut to mesenteric lymph nodes in a washout period after antibiotic cessation but *not during ABX treatment itself*. To our knowledge, whereas a handful of prior studies have documented antibiotic-induced inflammation^81^ the Knoop study is the only prior study reporting local and/or systemic inflammation which increases during the recolonization period after ABX cessation. The authors of that study concluded that this phenomenon could be an important contributor to adverse clinical outcomes after ABX exposure. We extend this finding here to report that post-ABX inflammation is diet-dependent with more severe ABX induced inflammation in animals randomized to AEN than in animals receiving PBEN.

Immune dysfunction during critical illness is a predictor of poor clinical outcomes. Several notable papers have detailed how ABX exposure can handicap the immune system, including a series of papers documenting the impact of AID upon hematopoiesis^51,82–86^. For example, Josefsdottir et al. demonstrated anemia, leukopenia, reduced bone marrow cellularity, and decreased splenic mass in a murine model of AID. Additional studies provided mechanistic details explaining how ABX disrupts a normally robust cross-talk between gut microbes and the bone marrow niche^82–84^. We reproduced many key findings from these prior studies, namely anemia, leukopenia, lymphopenia and a decrease in the B lymphocyte/T lymphocyte ratio in mice receiving broad-spectrum oral ABX. However, our data differs from this paper in one key regard. In our hands, circulating myeloid cells and pro-inflammatory cytokines increased markedly even as lymphocytes and total WBCs decreased in the context of ABX-induced inflammation.

These findings indicate that ABX not only suppresses overall bone marrow function but also favors myelopoiesis at the expense of lymphopoiesis. This is important because markers of myeloid skewing, e.g. increased neutrophil/lymphocyte ratio, have consistently been linked to increased mortality among hospitalized patients^52–55^. Importantly, we show ABX-induced bone marrow dysfunction to be diet-dependent with nearly all markers of BM dysfunction and myeloid skewing significantly exacerbated in animals receiving AEN. In most cases, animals randomized to receive PBEN after ABX displayed CBC and immune cell profiles similar to ABX-naive controls, indicative of recovery towards baseline. We confirmed that these diet-dependent immune changes have functional consequences relevant to patients receiving ABX with PBEN affording significant advantages in mouse models of nosocomial infection and endotoxemia. We repeatedly observed healthier animals with PBEN diet, manifest either by survival, systemic bacterial dissemination, or colonization levels in the gut. Together, these results provide compelling evidence that dietary interventions for ABX-treated patients could reduce the incidence of infectious complications after ABX treatment.

Finally, we conducted a small randomized single-center study of a nutritional intervention in critically ill children. In this study, we compared commercially available pediatric versions of PBEN and AEN in pediatric ICU patients requiring supplemental enteral nutrition. The study intervention itself did not involve a specific ABX regimen but all enrolled children received ABX during the study period. The study was not powered to identify differences in clinical outcomes (e.g. mortality or nosocomial infection). Rather, it was designed to compare diet-dependent changes in the gut microbiome as well as other laboratory values collected as part of routine clinical care. Consistent with the hematologic studies in mice, we observed significantly lower neutrophil/lymphocyte ratios in children receiving PBEN, despite similar total WBC and the acute phase protein C-reactive peptide. This provides further evidence that the PBEN diet mitigates myeloid skewing observed in the context of AID.

Despite a small sample size, the heterogeneous patient population, and the large number of confounding variables observed in ICU patients, we found clear diet-dependent differences in gut microbial diversity and taxonomic composition. Samples from children receiving PBEN were enriched common gut commensal anaerobes from the genera *Bacteroides*, *Phocaeicola*, and *Ruthenibacterium*. Although we did not conduct metabolomic analyses, CaZY analysis detailed the genomic capability of these organisms to enzymatically degrade dietary fiber, often producing SCFAs in the process. These microbial profiles differed dramatically from samples from children randomized to AEN which were enriched with *Streptococcus* species such as *S. salivarius*. *Streptococcus* is a diverse genus of facultative anaerobes with pathogenicity potential that is typically present in the oral cavity. Interestingly, enrichment of oral microbes such as *S. salivarius* in the distal intestine has been identified as a biomarker of generalized depletion of the gut microbiome after antibiotic treatment^87–89^. Our results will require validation in future studies, but they are biologically plausible and support published studies of dysbiosis evoked by antibiotics, added sugar, and emulsifiers in the diet with a corresponding lack of fiber.

We acknowledge that the human pilot study was small in scale and also acknowledge limitations of our animal studies. The animal model used does not fully reflect the complexity of the human ICU environment. Also, the liquid diets fed to mice are quite different from normal rodent chow, but we note that the same can be said for human ICU patients, i.e. commercial enteral formulas are distinct from standard human meals. The aggressive ABX regimen used in the study is routinely used in experimental model of antibiotic-induced dysbiosis. However, human patients often face similarly aggressive antibiotic exposures that are typically intravenous rather than enteral. Prior rodent studies of intravenous and intraperitoneal ABX as well as more limited oral ABX regimens have demonstrated similar patterns of AID and antibiotic-induced immune dysfunction, but future studies will need to determine how the route of ABX administration impacts these results.

In summary, our findings provide compelling evidence that all-natural plant based enteral nutrition can favorably shape the gut microbiome’s response to antibiotic exposure, immune function, and resilience to infections. These results have the potential to change the approach to the care and treatment of critically ill patients, nearly all of whom have at least some component of AID. Maintaining or re-establishing a diverse gut microbiome rich in beneficial microbes likely can improve clinical outcomes for such patients, although this has not yet been demonstrated in an adequately powered clinical trial.

## Data Availability

Sequencing data has been deposited and made publicly available as of the date of publication under bioproject number PRJNA1200130. All original code will be deposited and made publicly available as of the date of publication. Any additional information required to reanalyze the data reported in this paper is available from the lead contact upon request.

## Lead Contact

Further information and requests for resources and reagents should be directed to and will be fulfilled by the lead contact, Michael Morowitz.

## Material availability

This study did not generate new unique reagents.

## Data and code availability

- Sequencing data has been deposited and made publicly available as of the date of publication under bioproject number PRJNA1200130.
- All original code will be deposited and made publicly available as of the date of publication.
- Any additional information required to reanalyze the data reported in this paper is available from the lead contact upon request

## Acknowledgements

The authors express gratitude to the Flow Cytometry Core at UPMC Children’s Hospital for their invaluable assistance with this project. This research was partially support by the National Institute of General Medical Sciences of the National Institutes of Health (Award Numbers T32GM008208 and 1F30HL170777) and UPMC Children’s Hospital of Pittsburgh Research Advisory Committee Award. The content is solely the responsibility of the authors and does not necessarily represent the official views of the National Institutes of Health or other funding agencies. Functional Formularies provided some Liquid Hope for use in this study. The company was not involved in the planning, execution, or analysis of the experiments nor did it affect the authors’ conclusions.

## Author Contributions

Conceptualization, MC, JT, MJM; Methodology, MC, JT, MJM, MH,; Software, MR, JA; Investigation, MC, JT, RGR, GW, FF, RN, LC, AS, BF, JA, FF; Writing – Original Draft, MC, MJM; Writing – Review & Editing, RN, JA, BF, FF; Funding Acquisition, BC, DS, MJM; Resources, BF, MJM; Visualization, MC; Supervision, MJM

## Supplementary

**S1.**
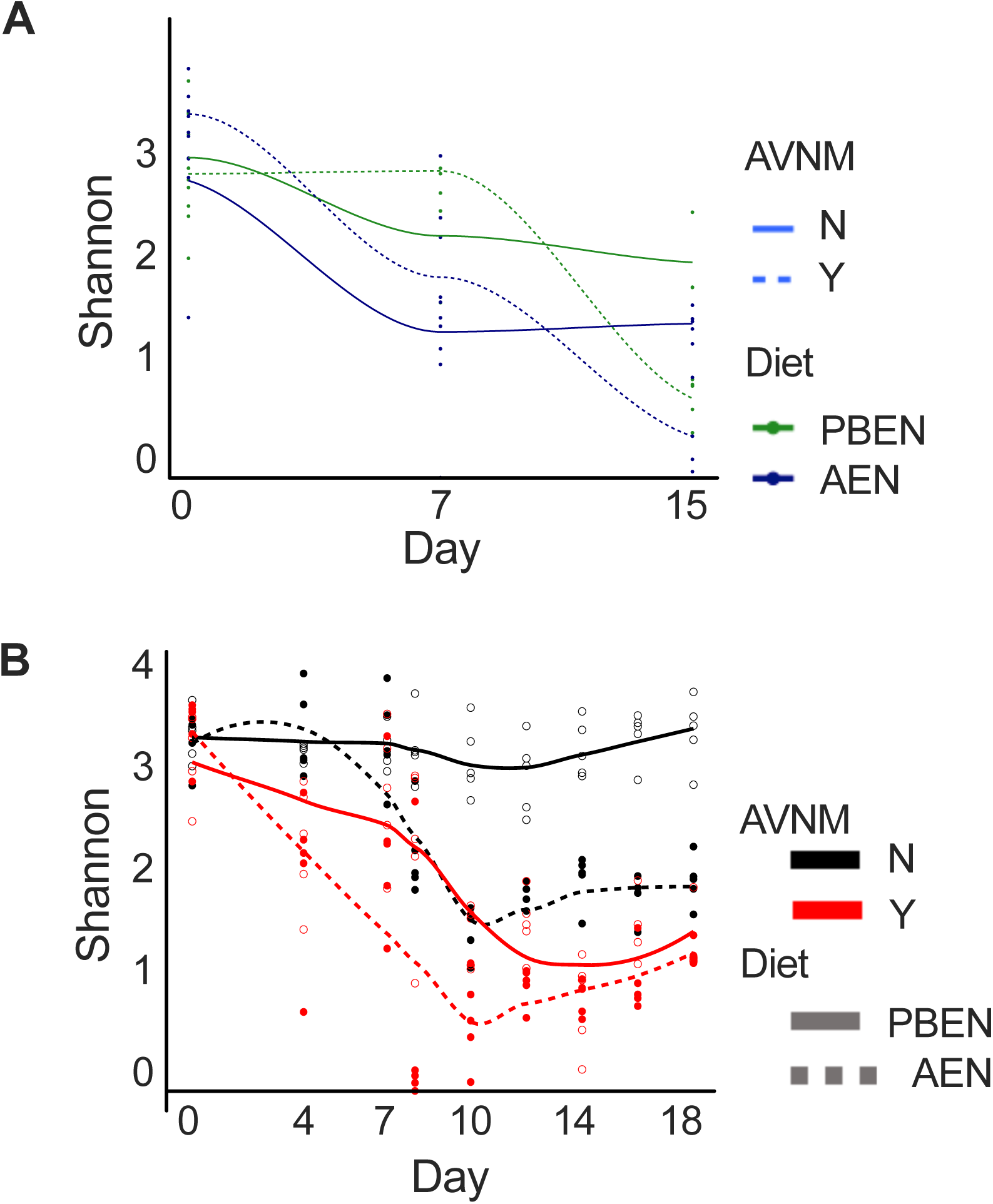
PBEN mitigates antibiotic-induced dysbiosis in different sequence iterations of ABX and diet treatments. Alpha diversity analysis of stool microbiome diversity in mice that A. received AEN during the initial week of antibiotics and were then randomized to PBEN or AEN (n=4 mice/diet) B. received AVNM in respective diets then continued on diets without ABX for an additional ten days (n=5 mice/diet).

**S2.**
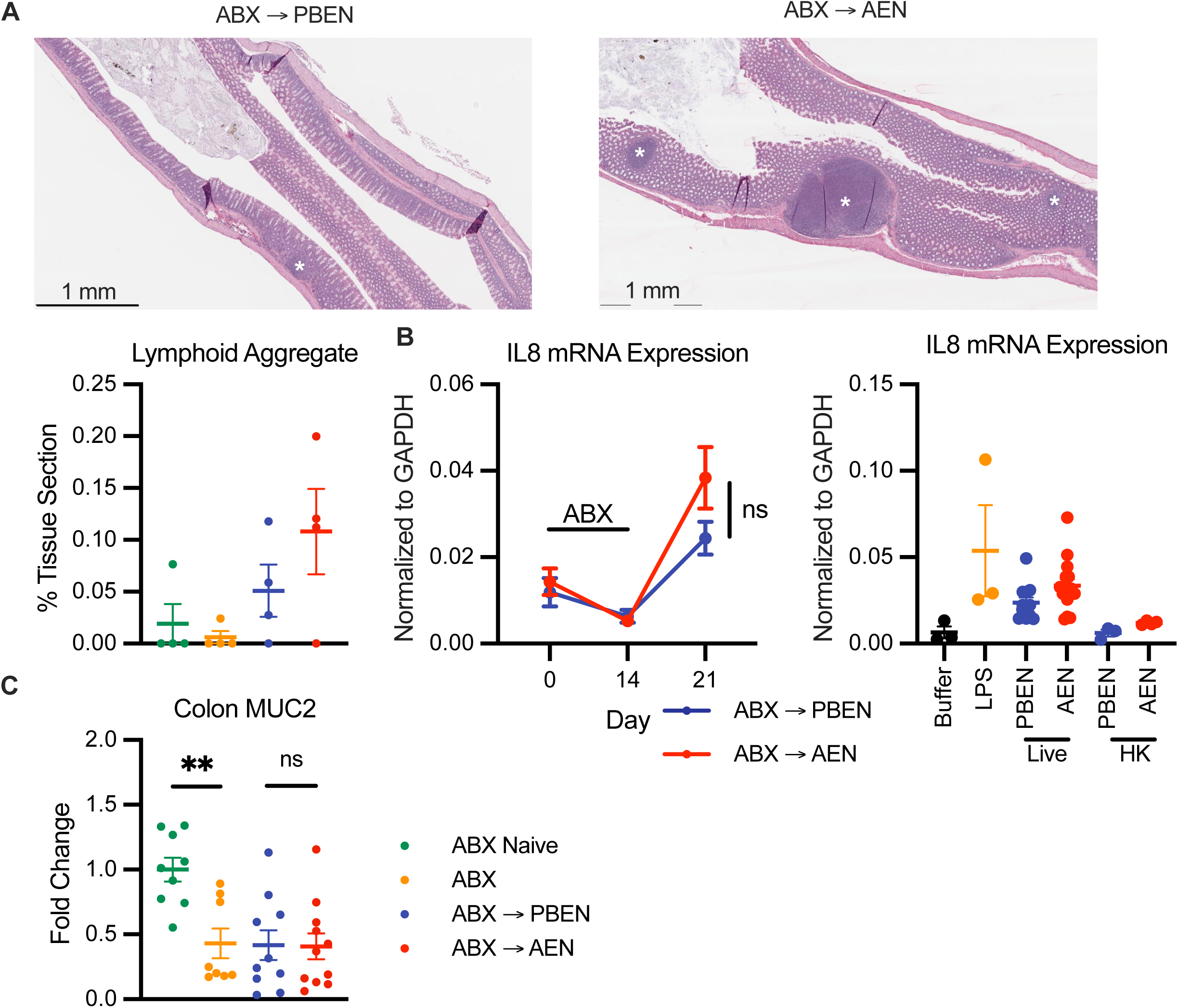
Antibiotic and diet induced changes in Mucin gene expression and colonic morphology. A. Representative H&E stained sections of colonic tissue in animals randomized to PBEN or AEN after two weeks of ABX with lymphoid aggregates marked with asterisks. Quantification of proportion of tissue occupied by lymphoid aggregate standardized to overall length of tissue was quantified in 4 sections per group. B. Quantitative RT-PCR analysis of IL-8 expression in HT29 colonic epithelial cells co-cultured with stool collected throughout ABX and diet treatments and with live or heat killed bacteria derived from fecal samples. C. Colon mucin-2 (MUC2) mRNA expression (n=8-11/group). All data represent mean ± standard error of the mean (SEM) with points representing individual mice. Student t test between PBEN versus AEN randomized animals from at least two independent experiments. ns, not significant; \**P* < .05, \*\**P* < .01, \*\*\**P* < .001, \*\*\*\**P* < .0001.

**S3.**
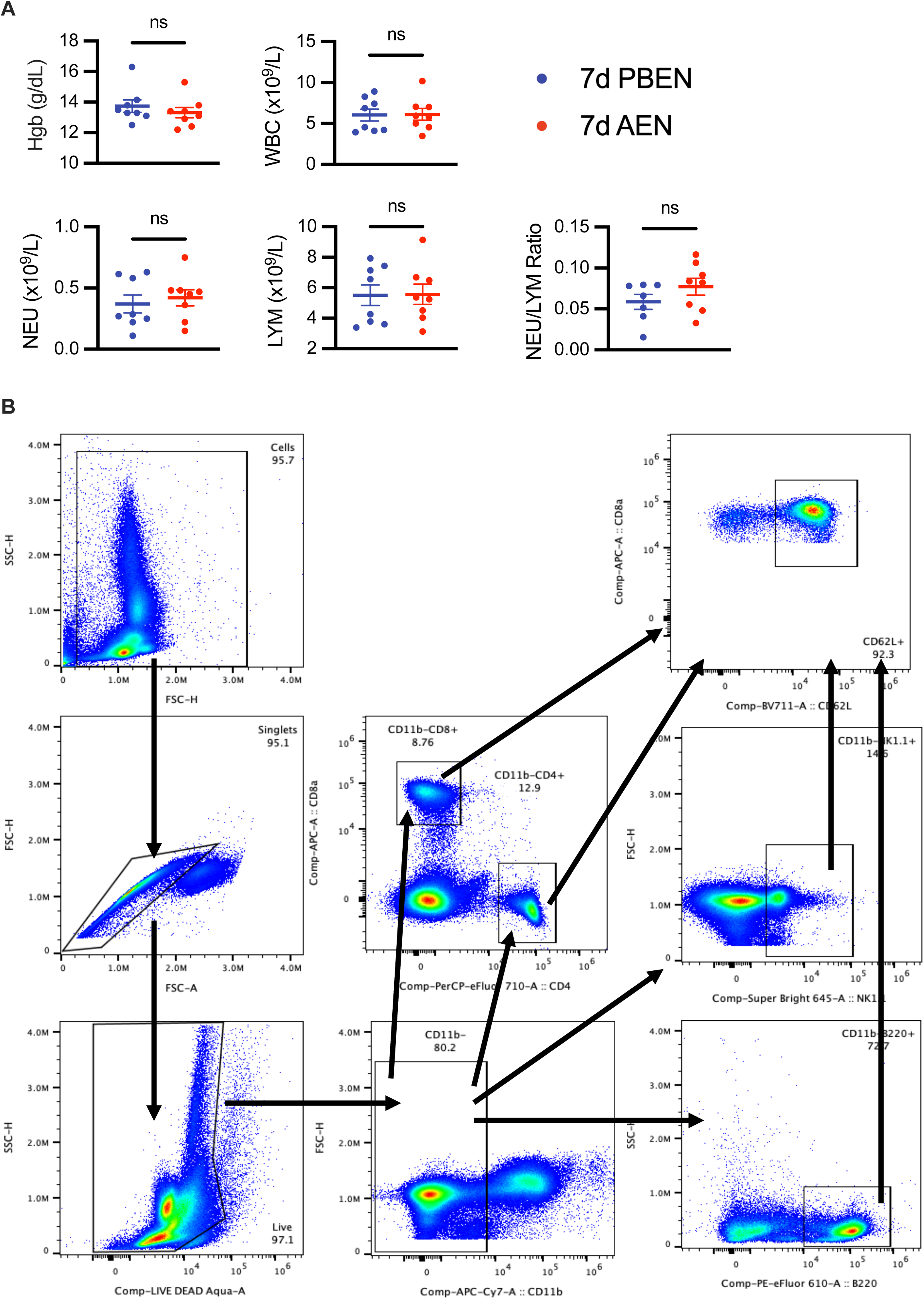
PBEN is superior to AEN in rescuing ABX-induced anemia and leukopenia. A. CBC profile of ABX naïve mice fed PBEN or AEN for seven days including red blood cells (RBC), white blood cells (WBC), neutrophils (NE), lymphocytes (LYM), and the neutrophil to lymphocyte ratio (NEU/LYM). B. Representative flow plot for white blood cell subsets. Debris was first filtered out using side-scatter (SSC) and forward scatter (FSC) followed by gating of live cells using Live Dead Aqua. CD11b-nucleated peripheral blood cell were divided to analyze T lymphocytes (CD4+ and CD8+), B cells (B220+), and NK cells (NK1.1+). Each of these populations was further analyzed for expression of CD62L. All data represent mean ± standard error of the mean (SEM) with points representing individual mice. Student t test between PBEN versus AEN randomized animals from at least two independent experiments. ns, not significant.

**S4.**
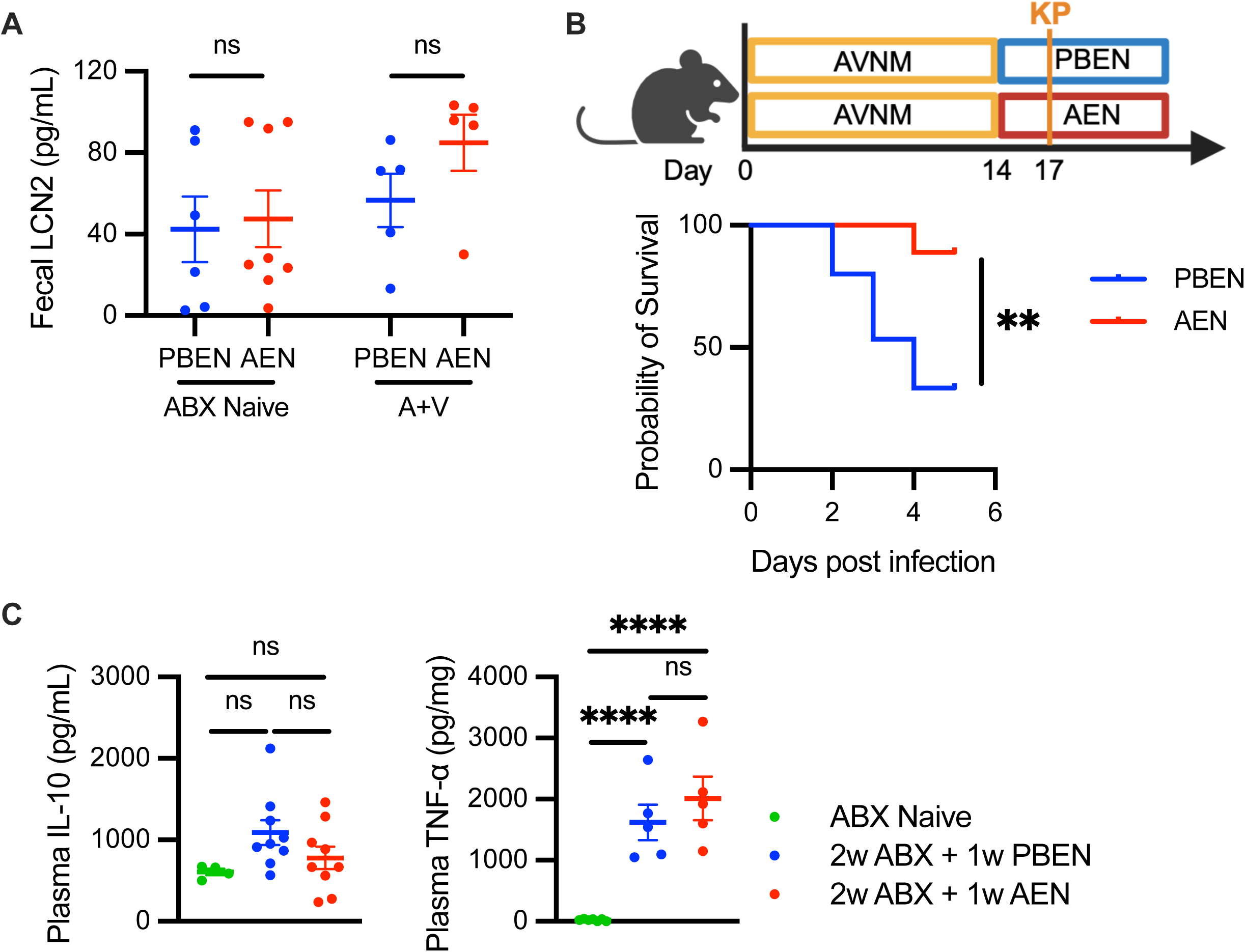
PBEN minimizes severity of post-antibiotic clearance of vancomycin-resistant *Enterococcus faecalis* and *Klebsiella Pneumoniae* GI infections but not *Klebsiella* induced pneumonia. A. Fecal lipocalin was measured in *Klebsiella Pneumoniae-*DSS experiment on day of sacrifice in experimental groups (n=5-8/group). B. Schematic experimental design of *K. Pneumoniae* (KP) induced pulmonary infection Mice received AVNM for two weeks and randomized to receive either PBEN or AEN. On the third day of washout animals were intratracheally inoculated with *Klebsiella Pneumoniae* 396 (KP396) and survival was monitored for six days. Kaplan-Meier survival curve comparing PBEN and AEN survival shown. C. Plasma IL10 and TNFa in mice receiving a sublethal dose of LPS and scarified after two hours. Survival following KP induced pulmonary infection analyzed using the Log-rank (Mantel-Cox) statistical test. Ordinary one-way ANOVA was used to compare the untreated control group and LPS-treated PBEN- and AEN-randomized animals and represented as mean ± standard error of the mean (SEM) with points representing individual mice. ns, not significant, \**P* < .05, \*\*\**P* < .001, \*\*\*\**P* < .0001.

**S5.**
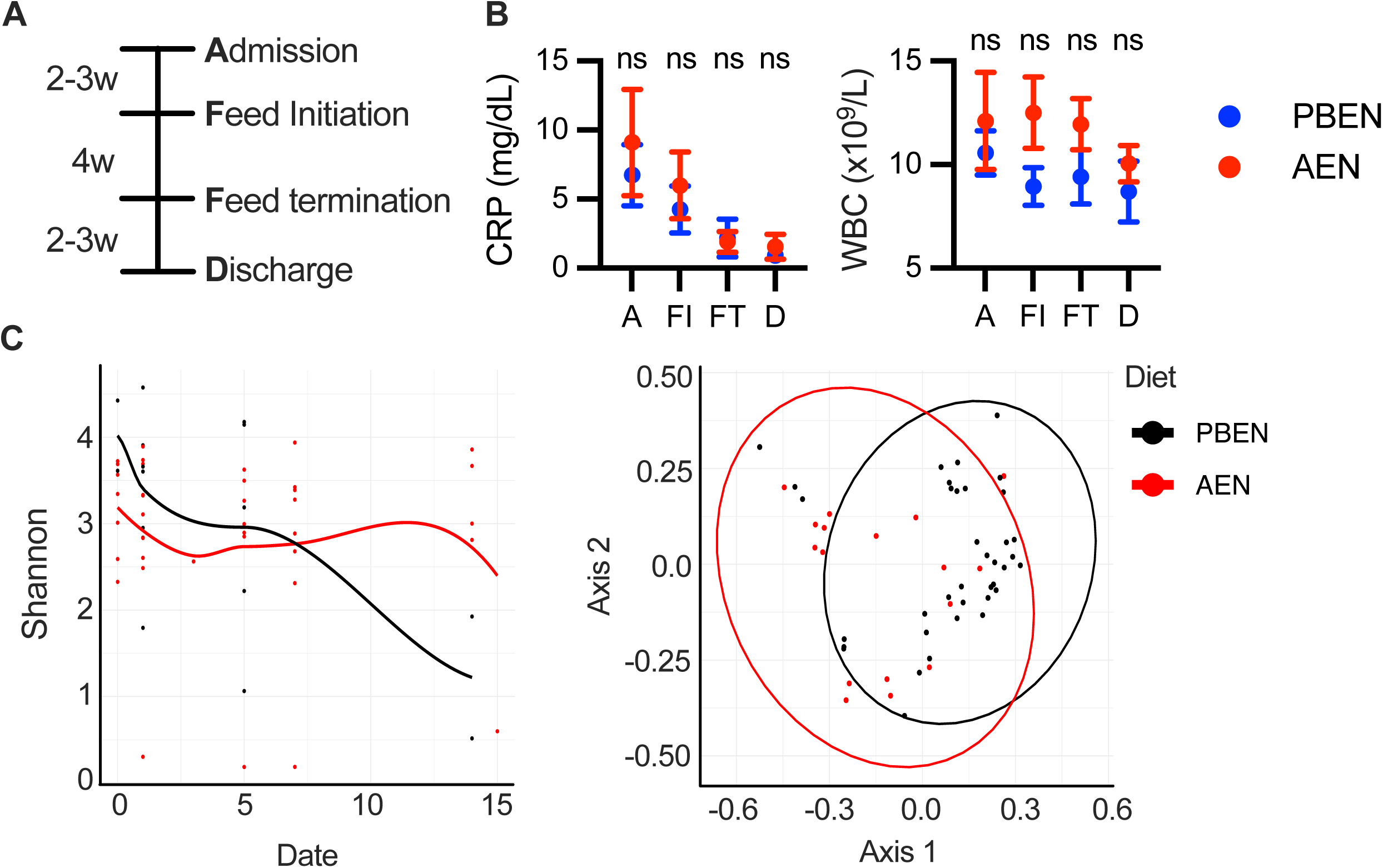
Dietary intervention with pediatric PBEN promotes microbiome recovery and improved peripheral blood profile in critically ill children. A. Schematic of intervention timeline. Blood was collected from patients at time of admission, feed initiation with either Nourish (PBEN) or Pediasure (AEN). Measurements of B. C-reactive protein (CRP) and white blood cells (WBC). C. Shannon diversity plot and unifrac analysis. All data represent mean ± standard error of the mean (SEM). Student t test between PBEN versus AEN randomized patients.

## STAR Methods

### EXPERIMENTAL MODEL AND SUBJECT DETAILS

#### Mice

Male C57BL/6 mice (8-10 weeks old, 20–30 g) obtained from Jackson Laboratories were used for all experiments. Mice were housed 3-4 mice/cage under a standard 12-hour light/ 12-hour dark cycle. All animals were randomly assigned to receive either standard chow, PBEN, or AEN with or without ABX consisting of ampicillin (1 g/L), vancomycin (0.5 g/L), neomycin (1 g/L), and metronidazole (1 g/L) supplemented with Splenda (4g/L). ABX were administered either in drinking water or in liquid diets. Liquid diets consisted of either standard adult polymeric formula, Vital; Abbott Laboratories, Lake Bluff, IL (AEN) and a plant-based whole foods formula, Liquid Hope; Functional Formularies, AEN West Chester, OH (PBEN). PBEN was diluted 4:5 with water to be isocaloric with AEN. All animal studies were approved by the University of Pittsburgh Institutional Animal Care and Use Committee.

#### Human samples

The study protocol for the clinical trial reported was approved by the University of Pittsburgh Institutional Review Board (Study 19080289, NCT 03414775). After obtaining written parental consent, 32 critically ill children admitted to the PICU of UPMC Children’s Hospital of Pittsburgh (Pittsburgh, PA, USA) were ultimately randomized in a 1:1 ratio to receive either commercially available pediatric PBEN (Nourish®, Functional Formularies, West Chester, OH, USA) or AEN (Pediasure®, Abbott Nutritional Products, Abbott Park, IL, USA). As outlined, three patients withdrew consent and were removed from the study resulting in a total of 13 patients randomized to Pediasure and 15 patients randomized to Nourish completing the study (Fig 7). Nourish was diluted 4:1 with sterile water to prevent feeding tube obstruction and contained 26.7 kcal/oz, 11.2g protein, 5.6g dietary fiber, 13.6g fat, and 6.4g total sugar per 8oz. Pediasure contained 30.3kcal/oz, 7g protein, <1g dietary fiber, 9g fat, and 12g total sugar per 8oz. Determination of patient ‘readiness’ for enteral feeds as well as route and rate of administration were at the discretion of the treating team. The time elapsed between admission and initiation of enteral feeds was determined exclusively by the clinical team without input from the research study team. Similarly, ‘readiness’ for discontinuation of enteral feeds was determined by the treating team. Nutritional consultation was obtained to ensure adequacy of dietary intake with both formulas. Exclusion criteria included: vasopressor use, history of allergy or intolerance to either formula, whey protein allergy or intolerance, gluten sensitivity or intolerance, or medical condition that necessitates the use of specific formula and/or nutritional needs (e.g. ketogenic diet for epilepsy).

Fecal samples and rectal swabs for analysis of the microbiome were collected on the day of feed initiation. For subjects remaining on enteral nutrition, additional samples were subsequently collected on day 5-7 and day 14 after feed initiation. All samples were obtained with a BD BBL Culture Swab as previously described^64^. If a subject failed to produce a fecal sample, then a rectal swab was collected. Samples were frozen at −80°C for batch processing and analysis. Data including demographic information, length of stay, positive culture, study diet utilization, and results of tests done as part of standard practice (e.g. complete blood count (CBC)) was extracted from the medical record and entered into a database. Antibiotic coverage was determined on a scale of 0-4 as coverage of anaerobic gram positive, aerobic gram positive, anaerobic gram negative, and aerobic gram negative bacteria. The study diet goal rate * hours was calculated as (actual volume of study diet administered / goal volume) * 24 to determine the amount of study diet received normalized to the goal rate set by the treating physicians.

The study was designed to enroll patients by convenience sampling between the predetermined time period between February 2018 and September 2019. Based on prior admission patterns, we estimated in advance that we would enroll up to 40 patients during this time window. Our previous laboratory studies and clinical trials indicated that this would provide adequate power to compare the abundance of gut microbes in patients receiving Nourish (PBEN) or Pediasure (AEN). Specifically, in a prior pilot study of eleven children reliant on enteral nutrition who transitioned from an AEN to PBEN diet^61^, we assessed microbial diversity at baseline and again after the transition. This approach allowed us to detect significant microbiome changes using LEfSe, a method based on the Kruskal-Wallis test that identified significantly enriched or depleted bacterial taxa. In the current study, we applied a similar approach, collecting baseline samples and additional samples after dietary interventions were introduced to monitor microbiome changes, our primary outcome.

The study was not powered to look at laboratory values such as CBC changes, or other clinical data such as mortality. However, we did record and monitor relevant laboratory values and medications. During the enrollment period, we randomized a total of 32 children to one of the interventional diets. One randomized patient improved before feed initiation and didn’t receive interventional diets, two consented patients were withdrawn from the study by parental request, and one patient was enrolled but samples were not available for analysis. Thus, 28 randomized patients (Table 1) initiated the interventional study. For the secondary outcome of CBC values, we excluded two patients identified as outliers using both ROUT and Grubbs’ analyses. One patient had a newly diagnosed leukemia, while the other exhibited a significant stress response to cannulation and decannulation from ECMO. Both primary outcomes (changes to the gut microbiome) and secondary outcomes (hematology values) are reported in this study. No harm occurred to patients because of participation in this study.

### METHOD DETAILS

#### 16s rRNA Gene Sequencing and Analysis

DNA extraction and 16S rRNA Gene Sequencing were performed at either Microbiome Insights (Richmond, BC, Canada) or Wright Labs (Huntingdon, PA, USA). DNA was extracted from samples using the Qiagen PowerSoil kit (Qiagen, Germantown, Maryland, USA) according to the manufacturer’s protocol. Amplicon PCR of the V3-V4 region (515F and 806R primers) of the 16S rRNA gene was performed on the extracted DNA. Sequencing was performed on the Illumina MiSeq platform utilizing the manufacturer’s v2 chemistry with paired-end 250 base pair reads.

Sequence data were analyzed by using Quantitative Insights into Microbial Ecology 2 (QIIME2)^90^. An ASV table was generated using the DADA2 pipeline after trimming 22 base pairs from the 5’ end of both forward and reverse reads. Taxonomy was assigned to reads using the QIIME2 naïve-bayes classifier trained on the 515F and 806R 16S amplicon from the Greengenes taxonomic database (version 13.5)^91^. A phylogenetic tree was generated from aligned ASV sequences using FastTree^92^. Finally, ASV table, taxonomic assignment, ASV sequences and tree were exported in hd5, tsv, fasta and newick formats respectively. Taxonomic assignments and metadata were combined with the ASV table into a single biom object using biom tools. This biom file along with the tree of ASVs were imported into R as a phyloseq object using the R phyloseq^93^ library.

For alpha and beta diversity analyses samples were rarefied to an even depth. Alpha and beta diversity were calculated using the R libraries phyloseq and vegan^94^. Alpha diversity was measured using both Chao1 and Shannon diversity metrics. Significant differences in alpha diversity between groups were compared using the pairwise Wilcoxon tests in R. Beta-diversity distances were measured using both the weighted-unifrac and bray-curtis metric. Variations in beta diversity between sample groups were assessed using the Adonis2 and PERMDISP algorithms in the Vegan R library. Differences in taxonomic abundances were compared using Analysis of Composition of Microbiomes 2 (ANCOM2)^95^.

#### Metagenomic Sequencing

DNA for metagenomic sequencing was extracted from stool and swab samples using the manual protocol for the Qiagen DNeasy 96 PowerSoil Pro QIAcube HT kit (Qiagen, Germantown, Maryland, USA). Library prep and metagenomic sequencing were performed at Wright Labs ((Huntingdon, PA, USA) on the Illumina HiSeq platform with paired-end 150 base pair reads.

Sequencing reads were passed through cutadapt^96^ to remove primer contamination, then trimmomatic ^97^ was used to remove low quality base pairs, finally reads were aligned against the human genome using the Burrows Wheeler Alignment (BWA) software^98^, and unmapped reads were returned.

On average 2.04 million reads per sample were obtained for further analysis (s.d 1.9 million reads). Taxonomic and functional profiles were generated by using Kraken2^99^ workflow, a metagenomic classification tool designed to classify sequences based on k-mer matches to Kraken2 Standard database (2022). These markers can be whole genomes, genes, or other markers. Kraken results were converted to a biom format otu table using the kraken2biom script, then the biom file was imported into phyloseq. Samples with less than 100,000 reads were removed from downstream analysis. Alpha diversity was calculated using only the Shannon entropy metric, since due to the nature of shallow metagenomic data, OTU abundance tends to be overestimated. Beta diversity distances were calculated using only the Bray-curtis metric as Unifrac methods require a phylogenetic, which would require a universal marker between OTUs.

#### Metagenomic Assembly

For each sample, an assembly of metagenomic reads were assembled using the megahit metagenomic assembler using a range of kmer sizes (21,29,39,59,79,99,119,141). Contigs from each assembly were assigned taxonomy using Kraken2. To estimate the abundance of each contig, BWA was used to map the sample reads back to the contigs, and Samtools^100^ was used produce depth of reads per contig. Abundance per contig was normalized to library size and contig length in an equivalent sense to fragments per kilobase per million bases (FPKM) occasionally used in transcriptomic experiments.

#### CAZyme Annotation

The CAZy gene encoding contigs from the metadata were identified and classified based on the CAZymes database^100^ by the carbohydrate-active enzyme analysis toolkit (CAT). A combination of DIAMOND^101^ and HMMER^102^ predictions were used to annotate contigs encoding CAZymes.

Putative plant cell wall polysaccharide-degrading enzymes belonging to different CAZy families were identified and classified based on sequence-based annotation. The CAZyme encoding contigs were analyzed manually for different classes of CAZymes: glycoside hydrolases(GHs), glycoside transferases(GTs), carboxyesterases(CEs), carboxybinding module family(CBMs), and polysaccharide lyases (PLs). Subsequently, the CAZy results obtained were analyzed manually to determine the proportions of the different CAZymes present in the rumen metagenome data.

Differential abundance testing of CAZYmes was performed on RPKM values using multiple Wilcoxon-tests. As many patients had only one sample we selected one representative sample from each patient. Where multiple samples were present we chose samples that occurred multiple days after the diet started

#### Complete blood counts

Peripheral blood samples were collected from mice in EDTA treated tubes via cardiac puncture and hematology analysis was performed with VetScan HM5 (Zoetis, USA).

#### Flow cytometry

Mouse bone marrow was obtained by flushing the left femur. Blood and bone marrow cells were processed to remove red blood cells and debris. Total cell counts and viability were assessed on cellometer (Nexcelom Bioscience). Antibody panels were used for surface staining and analyzed on Cytek Aurora flow cytometer (Cytek Biosciences) and data was analyzed using FlowJo Software.

#### Lipocalin and Reg3g ELISA

Fecal samples for lipocalin and ileum tissue for Reg3g were collected and snap frozen and stored at −80°C until further processing. Samples were homogenized in PBS containing protease and phosphatase inhibitors, and supernatant was used for ELISA according to manufacturer’s instructions. All ELISA assays were performed in duplicate, and the average values were used for data analysis.

#### Luminex

Feces was removed from colon tissue samples obtained from experimental animals and the colons were immediately snap-frozen in liquid nitrogen for storage at −80°C until further use. Frozen colon tissue samples were homogenized in RIPA lysis buffer and protein concentrations were in tissue lysates were determined using the BCA assay according to the manufacturer’s instructions. Equal amounts of total protein from each sample were then used for subsequent Luminex multiplex analysis and analyzed using MagPix multiplex instrument.

#### Coculture of stool with HT29 cell line

Stool samples were homogenized in PBS, and bacteria were isolated for coculture by homogenizing in PBS and filtering. The bacterial pellet was resuspended in PBS containing 25% glycerol for storage. Quantification of bacterial concentration was achieved by measuring the optical density (OD) of the suspension at 600 nm. To obtain a standardized inoculum for coculture experiments, the bacterial concentration was adjusted to an OD of 0.05. For heat-inactivated controls, an aliquot of the bacterial suspension was incubated at 70°C for 30 minutes. For coculture, HT29 cells were seeded at a density of 300,000 cells per well in a 12-well plate the night before coculture and incubated at 37°C overnight in RPMI media supplemented with 10% FBS. The next day, 60 μL of the standardized bacterial homogenate was added to the HT29 cell wells and then incubated for 6 hours. RNA was collected and processed for qPCR analysis.

#### RNA extraction and qPCR

RNA was isolated in QIAzol Lysis Reagent based on manufacturer protocol followed by assessment of RNA concentration and integrity using NanoDrop. Normalized RNA was transcribed to cDNA. Quantitative real-time PCR (qPCR) was performed using QuantStudio3 Real-Time PCR System (Thermo Fisher Scientific) with PowerUp SYBR Green Master Mix. Relative gene expression was calculated using the ΔΔCt method using housekeeping genes GAPDH or b-Actin as a reference for normalization.

#### Spleen Culture

Spleens were aseptically collected and weighed before snap freezing. They were stored at - 80°C until further use. On the day of culturing, spleens were thawed on ice. To break up the tissue and release bacteria spleens were first punctured using a sterile pipette tip then homogenized via vortexing with 1.4mm ceramic beads in 1mL/g of sterile PBS for ten minutes. Homogenate was serially diluted and plated on blood agars for 48 hours at 37°C.

#### LPS Inflammatory Challenge

Mice either received the standard AVNM cocktail in PBEN and AEN or in standard drinking water for two weeks followed by one week of recovery with respective diets without ABX. After the recovery period, all mice were administered an LPS inflammatory challenge using Escherichia coli O111:B4 diluted in phosphate buffered saline (PBS) and administered intraperitoneally (i.p.) at a sublethal dose of 4mg/kg or 10 mg/kg to monitor survival. Animals injected with sublethal dose were sacrificed two hours post injection. Plasma was obtained by centrifuging whole-blood then used for TNFa ELISA per manufacturer’s instructions. For animals in the survival experiments, mice were observed daily twice daily, and mortality was recorded for one week after the injection of LPS.

#### *Vancomycin Resistant Enterococcus* Infection

All experiments with VRE were conducted according to guidelines approved by the University of Pittsburgh IBC (protocol IBC201900106). An overnight culture of Vancomycin-resistant *Enterococcus faecium* strain 34-10-S (VRE) was grown to saturation in BHI medium with 10 µg/ml vancomycin. Inoculums of VRE were prepared for gavage by diluting to 5×10^6^ CFU/ml in sterile PBS. Mice were infected by gavage with 10^5^ CFUs of VRE after being exposed to 7 days of ampicillin (1g/L). Bacterial counts were determined by plating serial dilutions of stool samples on agar plates with vancomycin. VRE colonies were identified by their ability to grow on vancomycin treated bile esculin agar (BEA) plates.

#### *Klebsiella pneumoniae* intratracheal infection

Mice were treated with ABX for two weeks followed by three days of recovery with PBEN or AEN. On the third day, they were infected with a hypervirulent mucoid K1 serotype clinical isolate of *Klebsiella pneumoniae* (KP396) and continued on their respective diets for an additional seven days. KP396 was grown overnight at 37°C at 220 rpm in tryptic soy broth (TSB). On day of infection, the overnight culture was diluted to 8,000 CFU/L and 400 CFU was given per mouse via intratracheal administration.

#### Klebsiella pneumoniae-Colonized Model of DSS-Induced Colitis (KP-DSS)

Mice were randomly assigned to receive either PBEN or AEN for three days. Subsequently, half of the mice in each group were administered antibiotics consisting of vancomycin (2.5g/L) and neomycin (5g/L) for 72 hours. On Day 7, all mice were colonized with wild-type *Klebsiella pneumoniae* via gastric gavage at a dose of 1 × 10D colony-forming units (CFUs) per 0.1 mL per mouse. Colitis was induced on Day 19 by providing 4% dextran sulfate sodium (DSS) in the drinking water. After three days of DSS treatment (Day 22), mice were euthanized for tissue collection. Stool samples were collected throughout the experiment, and the spleen was aseptically removed on the day of sacrifice. Primers previously designed^103^ against KP396 were used to assess *Klebsiella pneumoniae* burden in stool and spleen through quantitative polymerase chain reaction (qPCR).

### QUANTIFICATION AND STATISTICAL ANALYSIS

Statistical analysis was performed using GraphPad Prism (GraphPad Software) and R. Data are presented as mean ± standard error mean (SEM) unless otherwise indicated. Demographic and laboratory data was analyzed by Student’s T test, Fisher’s exact test, or Mann-Whitney test as appropriate and specific methods are specified in each individual figure legend. Outlier analysis was done on CBC values for patients as specified above. Significance is defined as *P < 0.05, **P < 0.01, ***P < 0.001, and ****P<0.0001.

#### Key resources table

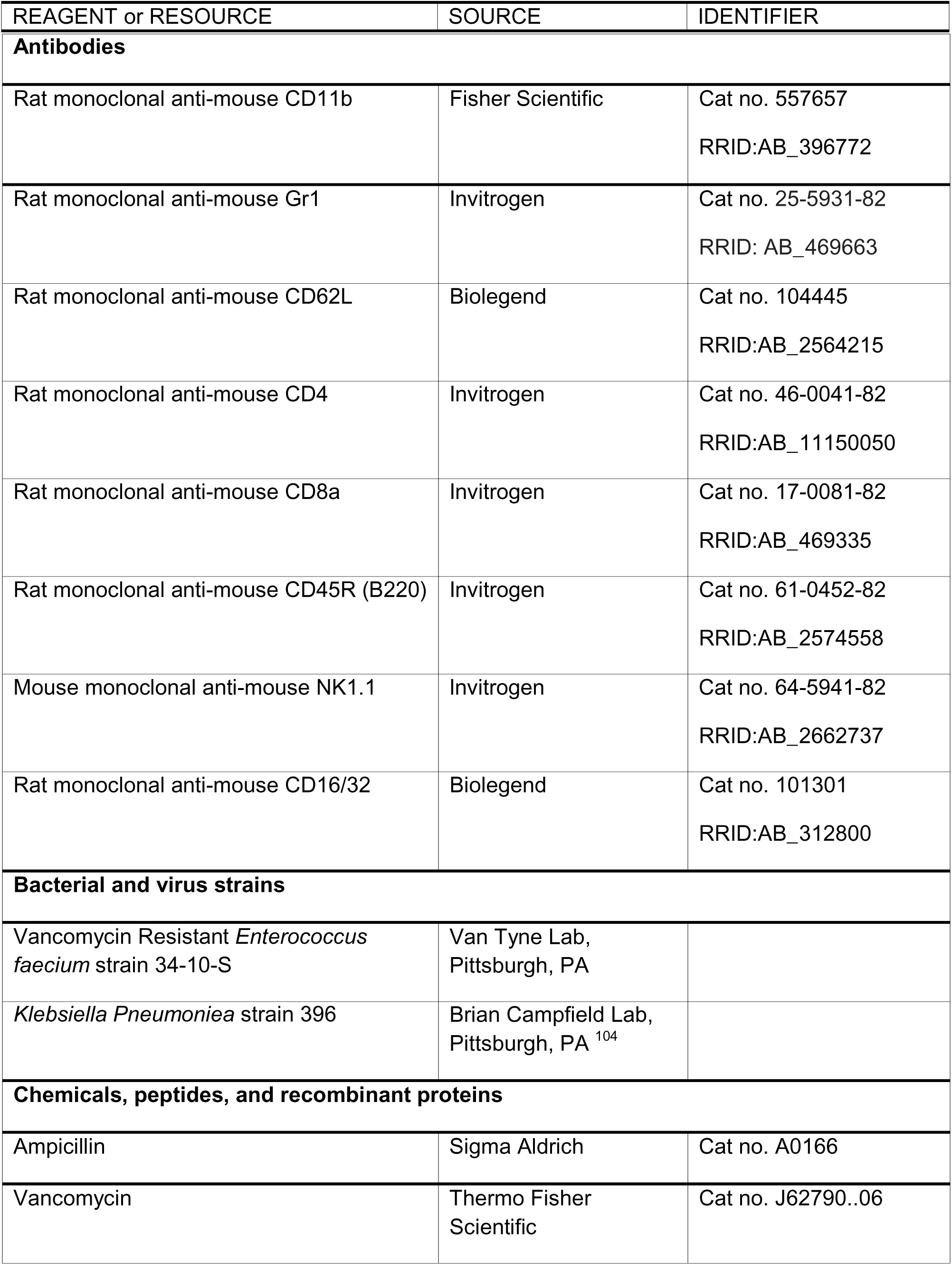
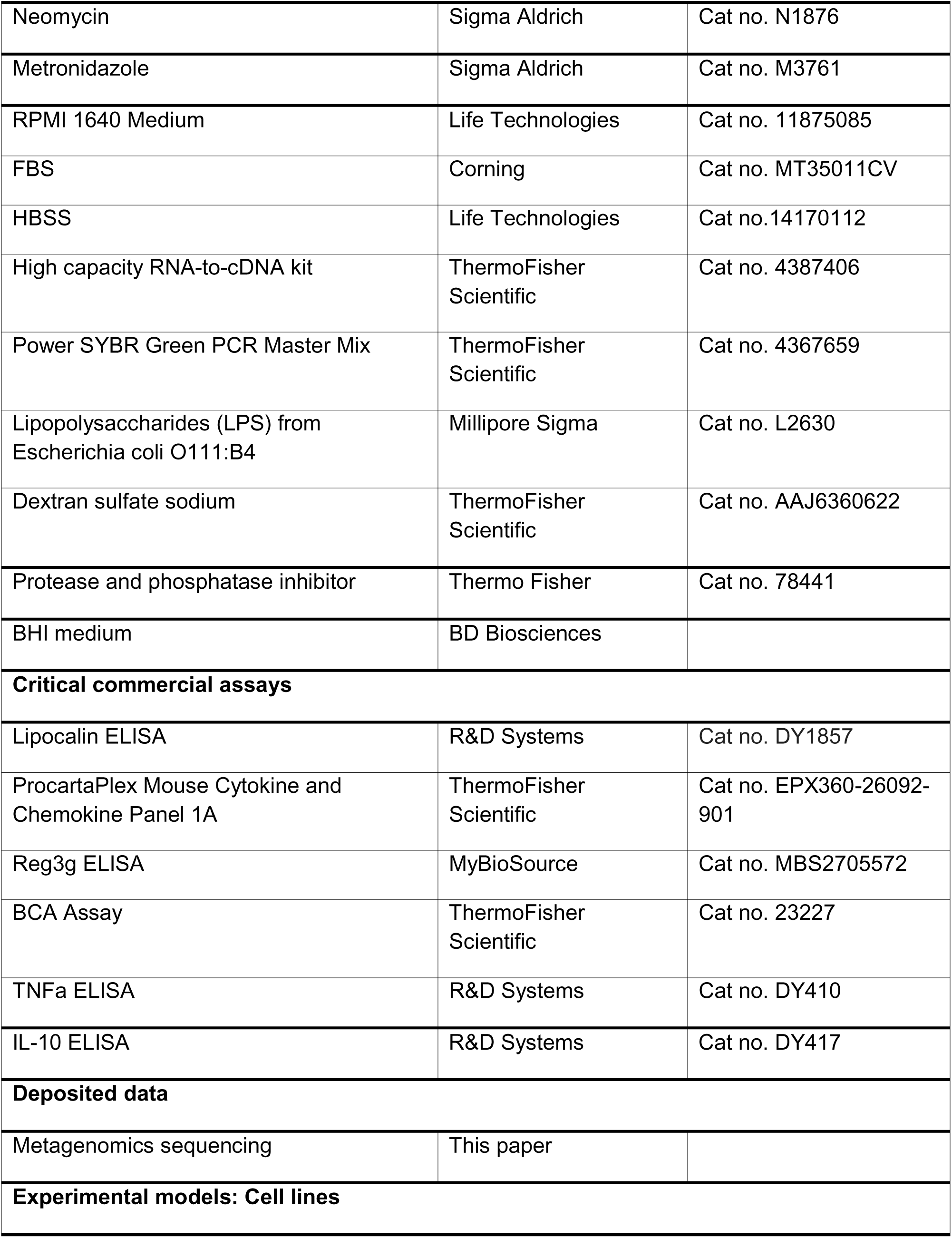
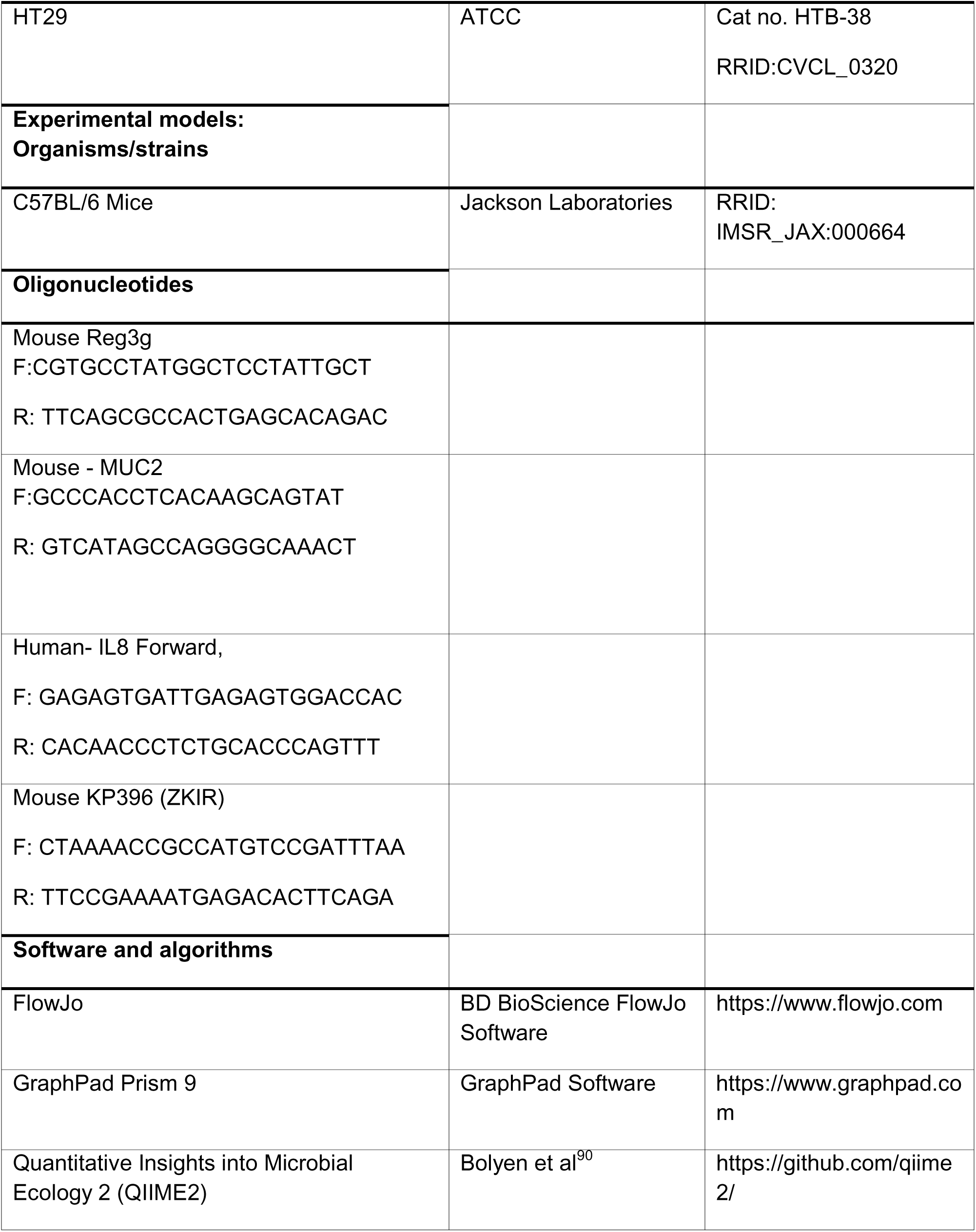
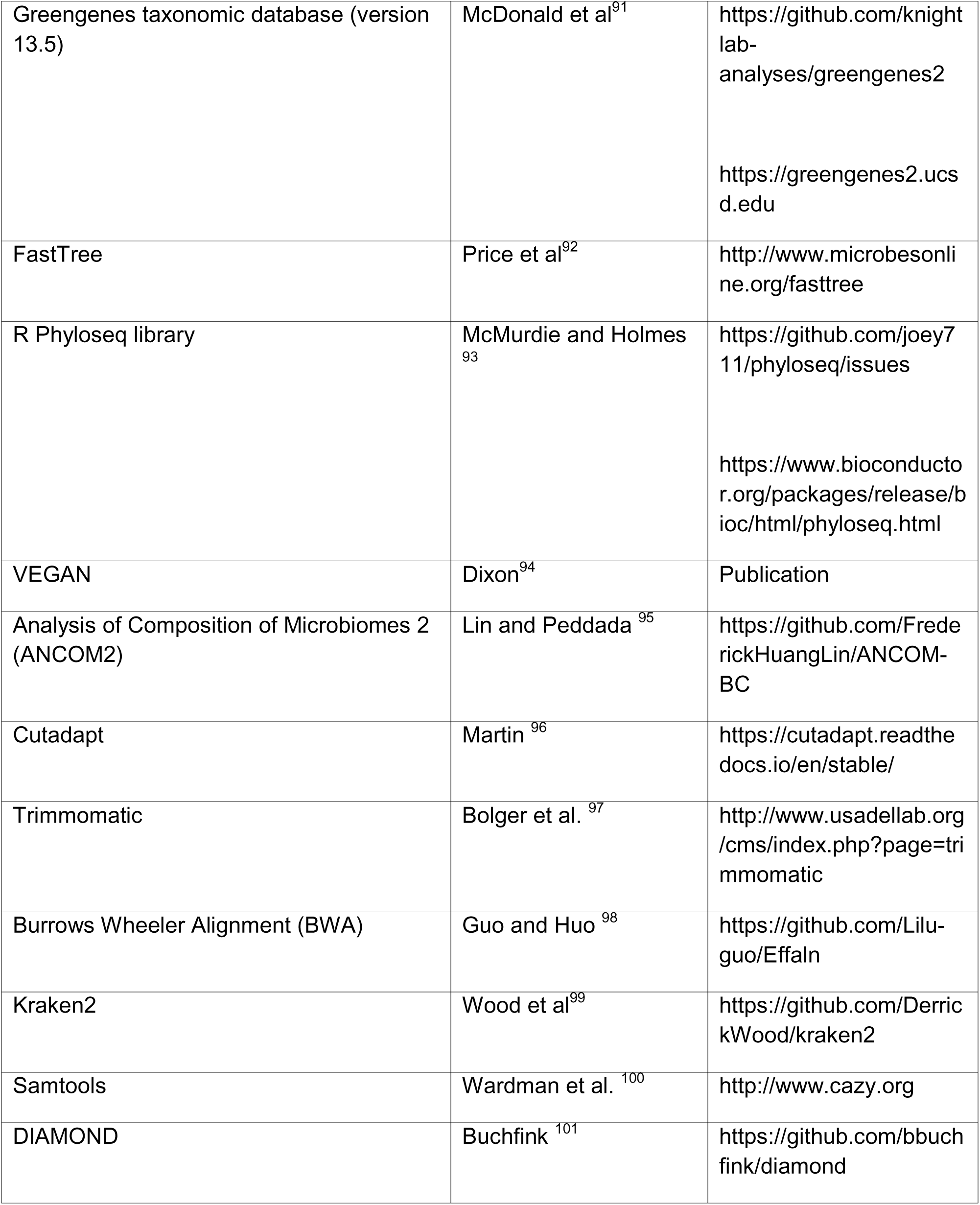
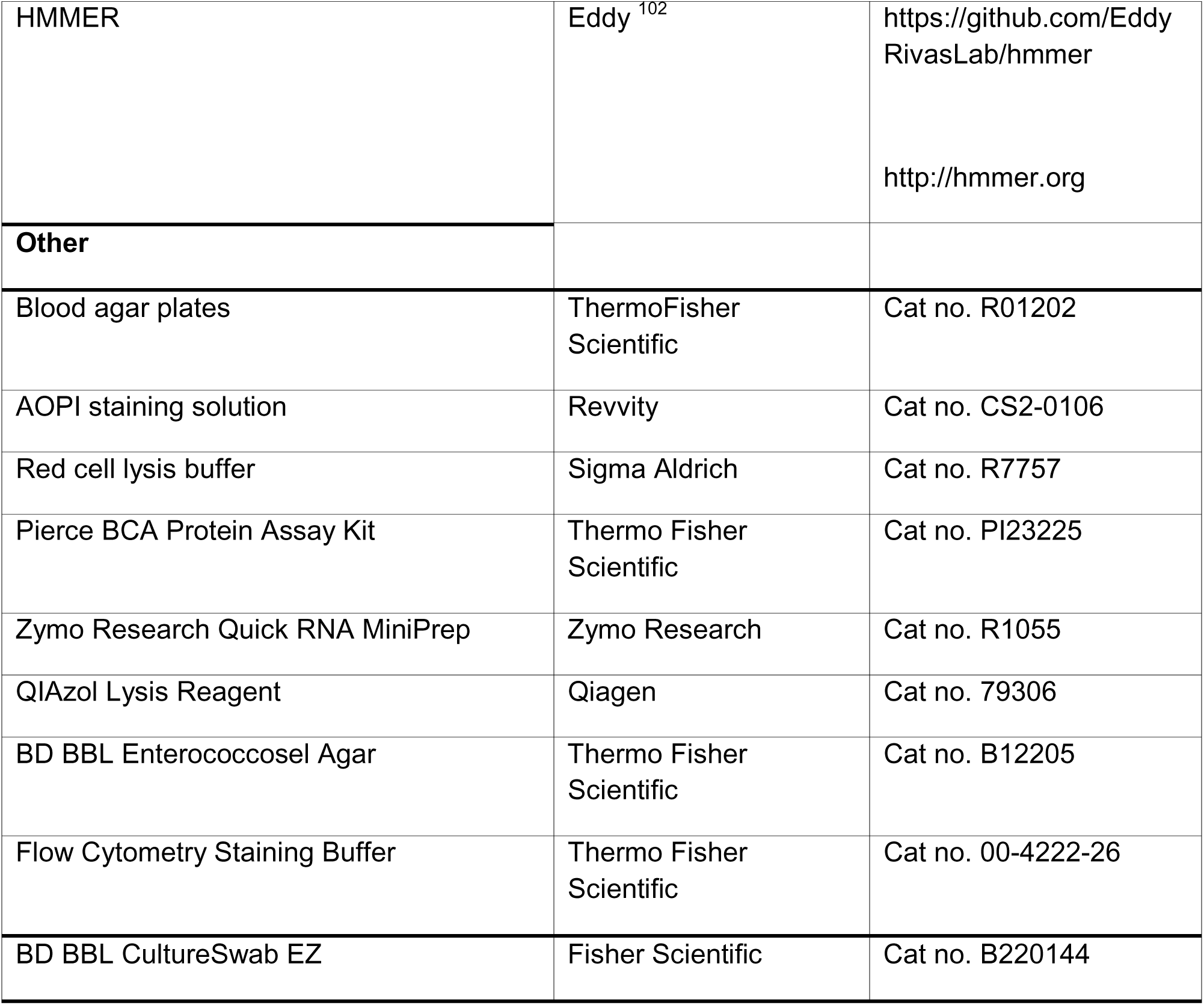

